# Factors confounding the athlete biological passport: a systematic narrative review

**DOI:** 10.1101/2021.03.26.21254386

**Authors:** Bastien Krumm, Raphael Faiss

## Abstract

**Background:** Through longitudinal, individual and adaptive monitoring of blood biomarkers, the haematological module of the athlete biological passport (ABP) has become a valuable tool in anti-doping efforts. The composition of blood as a vector of oxygen in the human body varies in athletes with the influence of multiple intrinsic (genetic) or extrinsic (training or environmental conditions) factors. In this context, it is fundamental to establish a comprehensive understanding of the various causes that may affect blood variables and thereby alter a fair interpretation of ABP profiles.

**Methods:** This literature review described the potential factors confounding the ABP to outline influencing factors altering haematological profiles acutely or chronically.

**Results:** Our investigation confirmed that natural variations in ABP variables appear relatively small, likely—at least in part—because of strong human homeostasis. Furthermore, the significant effects on haematological variations of environmental conditions (e.g. exposure to heat or hypoxia) remain debatable. The current ABP paradigm seems rather robust in view of the existing literature that aims to delineate adaptive individual limits. Nevertheless, its objective sensitivity may be further improved.

**Conclusions:** This narrative review contributes to disentangling the numerous confounding factors of the ABP to gather the available scientific evidence and help interpret individual athlete profiles.

**Key points:** Through longitudinal, individual and adaptive monitoring of blood biomarkers, the haematological module of the athlete biological passport (ABP) has become a valuable tool in anti-doping efforts.

This literature review described the potential factors confounding the ABP to outline influencing factors altering haematological profiles acutely or chronically.

While our results support the current ABP paradigm as rather robust to delineate adaptive individual limits, our work may contribute to disentangling the numerous confounding factors of the ABP to gather the available scientific

## 1 Introduction

The concept of an athlete biological passport (ABP) was developed in the early 2000s and officially introduced in 2009 [1], marking a turning point in the anti-doping field. With the haematological and steroidal modules of the ABP, blood and urine variables were no longer utilised exclusively as direct screening parameters but as robust markers of either erythropoietic stimulation or steroid intake in individual, longitudinal monitoring [2]. This indirect screening approach is pertinent because indirect biological markers may reveal abnormal variations in blood and urine potentially induced by doping [3].

The ABP is implemented with an adaptive probabilistic model based on a Bayesian approach [4] to determine the probability that an athlete’s haematological variations are of prohibited origin [5]. Therefore, the effect of doping is not directly examined via the modification of the biological parameters but via the variation in the mean and standard deviation of these biomarkers [3]. When a first sample is collected, upper and lower thresholds are determined with population-based average benchmarks [5]. These individual limits are then subsequently and progressively adapted based on each athlete’s values as additional samples are taken [5].

Fourteen parameters are currently recorded and analysed as part of the ABP haematological module in the online Anti-Doping Administration & Management System (ADAMS), which was developed and supported by the World Anti-Doping Agency (WADA). In ADAMS, two primary markers are monitored with remarkably strict operating guidelines [1]: haemoglobin concentration ([Hb]) and the erythropoiesis stimulation index ‘OFF-Score’ (OFFs), calculated as OFFs = [Hb] − 60 × √Ret%, including the percentage of reticulocytes (Ret%) and [Hb] in g.L^-1^. An atypical passport finding (ATPF) is then a ‘result generated by the adaptive model in ADAMS which identifies either a primary marker value outside the athlete’s intra-individual range or a longitudinal profile of primary marker values (sequence deviations) outside expected ranges, assuming a normal physiological condition’ [1]. A level of specificity of 99% (outliers correspond to the values outside of the 99%-range i.e. at least 1:100 chance that this result is due to natural physiological origin) is required for the system to notify an ATPF.

The ABP uses this quantitative Bayesian model to detect an ATPF. Then, a qualitative expert review by the Athlete Passport Management Unit (APMU) handles administration of the individual passport before eventually requesting an evaluation by three independent ABP experts [4].

The latter expert review is essential because factors other than doping, such as physiological variations of biological origin or the results of an athlete’s activity [3], may explain variations in haematological biomarkers. Interestingly, the plasma volume (PV) is at the core of many haematological changes [6], and [Hb], which is, by definition, measured as a concentration, may significantly vary and become difficult to interpret if PV changes [7]. In an athletic context, red blood cells diluted in plasma will be affected differently by conditions such as acute and chronic exercise, environmental factors or certain illnesses [8], and these conditions can be defined as confounding factors leading to a potential misinterpretation of the ABP biomarker variations [9].

The variability caused by confounders can, however, be significantly reduced by understanding the nature of these factors [3]. Four main sources of variation may be outlined: pre-analytical and analytical conditions, physical exercise, environmental conditions and individual characteristics. Currently, very strict guidelines for blood collection and analyses [10,11] address the effect of pre-analytical and analytical variations. In the current ABP operating guidelines [1], the notion of confounding factors is outlined only for the steroidal module (i.e. urine testing) of the ABP with reference to a recent review of confounders by Kuuranne et al. (2014) [12]. However, to the best of our knowledge, no systematic review has thoroughly examined the potential confounders affecting biomarkers of the haematological module. We hypothesise that the current ABP approach may be challenged by the scientific evidence of significant confounders and that a thorough review of the latter may help experts in their interpretation of abnormal ABP profiles.

Therefore, this systematic review investigates the existing literature regarding the various factors influencing haematological markers. Its results, presented in a narrative format, aim to enhance anti-doping actors’ prevailing understanding of these confounders.

## 2 Methodological approach

Nine potential confounding factors of the haematological module were identified at the beginning of this review: doping practises, acute exercise, chronic training, exposure to a hot environment, exposure to a cold environment, exposure to a hypoxic environment, individual disorders or diseases, athlete characteristics and pre-analytical factors (Figure 1).

**Figure 1:**
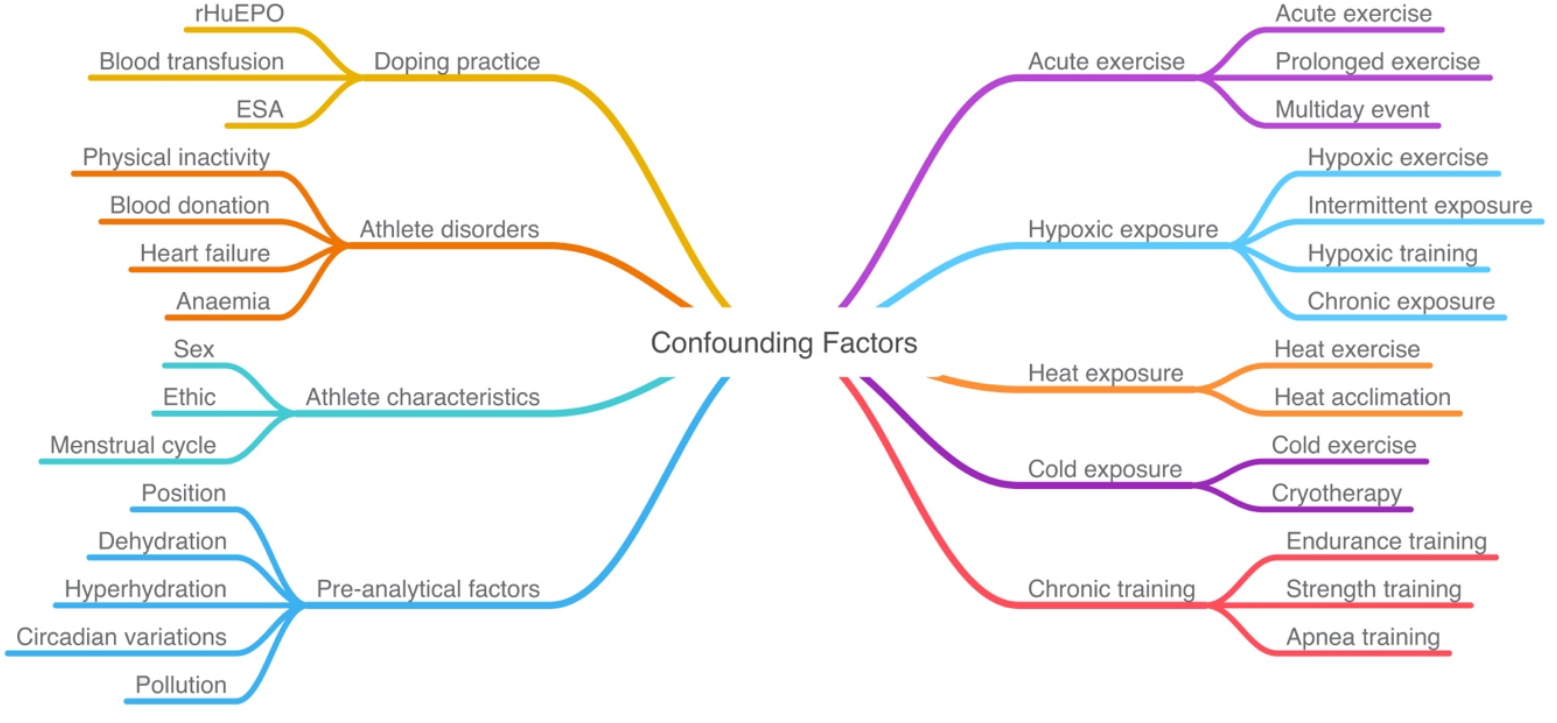
Illustration of initially identified confounding factors.

A literature search was conducted in June 2020 on the PubMed and Google Scholar platforms with the above factors as research terms used in various combinations with the following terms: (haemoglobin OR haematocrit OR reticulocyte OR plasma volume) AND (haematology OR haematology OR haematological variation OR haematological parameter) AND (athlete) AND (sport). From the results, the authors selected the most relevant articles considering the broad spectrum of different confounding factors potentially involved in blood variations. The main findings are presented in the result tables. Initially, all resulting titles and abstracts (n = 2325) were screened with adequate articles retained for further evaluation (n = 535). Grey literature was included by screening the references of the most relevant articles. Only studies in English with human subjects were considered for inclusion.

Studies where the effects of at least two confounding factors could not be clearly discriminated were excluded. Studies including both of the two primary ABP biomarkers (i.e. [Hb] and Ret%) were identified in a first selection round (n = 125). At this stage, however, no studies related to climatic conditions (heat and cold exposure) matched the latter inclusion criteria, despite the fact that these conditions exhibit an impact on PV variations affecting ABP markers. Hence, several articles related to environmental conditions but reporting no direct measurements of Ret% or [Hb] were included in this review when PV changes (with a subsequent impact on [Hb]) were deemed pertinent. The process resulted in the selection of 83 pertinent studies in an anti-doping context where added value for the interpretation of ABP profiles was identified (as illustrated in the PRISMA flow diagram below in Figure 2).

**Figure 2:**
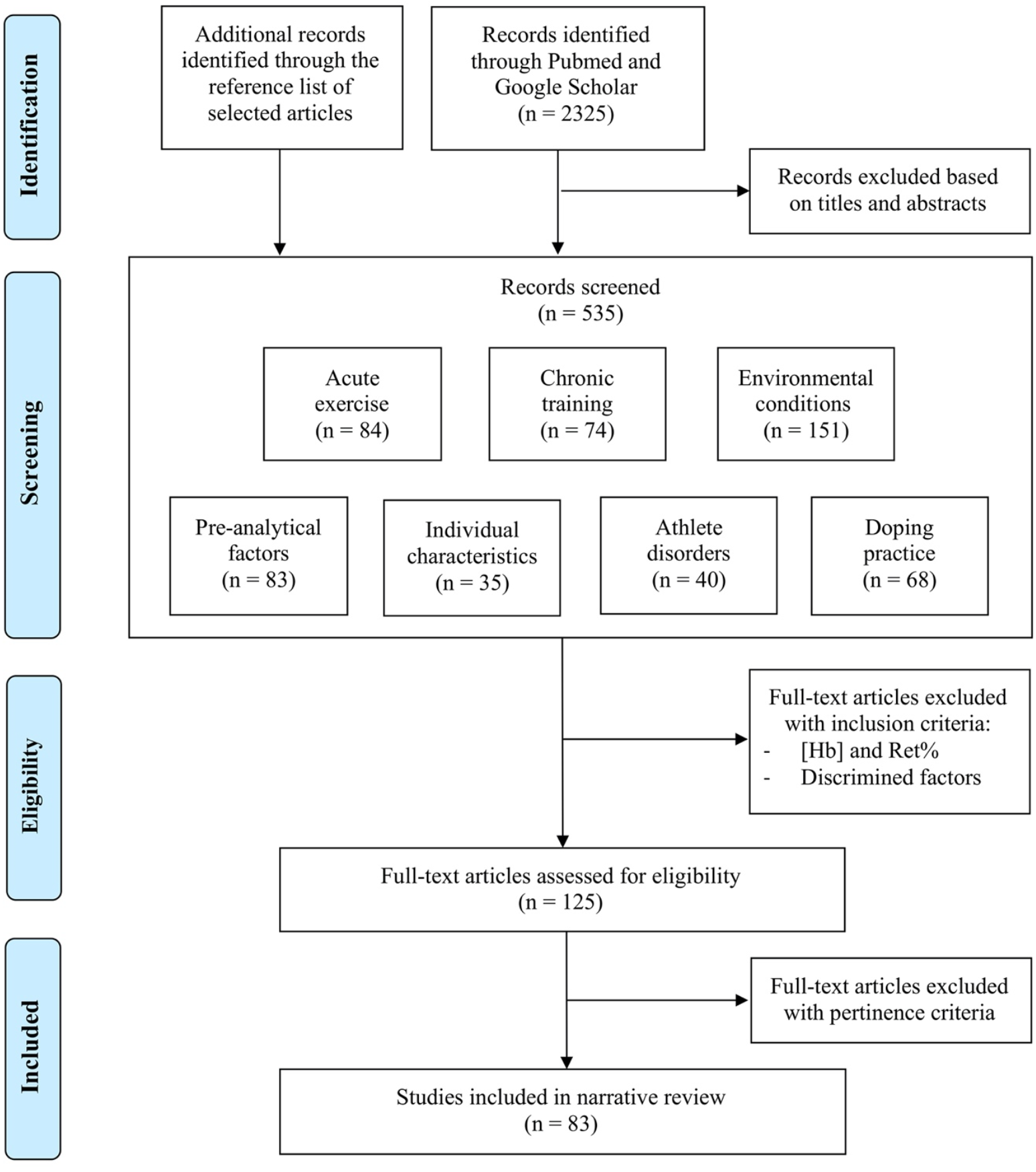
“PRISMA” flow diagram (Moher et al., 2009) Haemoglobin concentration ([Hb]); reticulocytes percentage (Ret%).

Among the multiple causes identified, some pre-analytical factors, such as acute exercise or exposure to various extreme environments, may exert a significant influence on the haematological biomarkers. The period of influence these factors had ranged from a few minutes (e.g. body position during sampling [13]) to a few hours (e.g. intense exercise [14]), and can, therefore, alter blood variables. With the existence of strict WADA guidelines ruling out confounders mentioned in the doping control forms [1], this narrative review will, however, focus mainly on confounding effects not addressed by the latter WADA rules. For this review, we have considered the relative changes observed after an intervention or in a specific condition for the following physiological variables: [Hb], Ret%, OFF-Score, haematocrit (Hct) and PV.

## 3 Results of the literature search

The literature review indicates that doping practises represent a major confounder altering the ABP haematological variables most significantly (Tables 1 and 2). Other significant changes were observed after prolonged exercise (Tables 5 and 6), exercise training (Table 7), training periodisation (Table 8), thermal acclimation (Table 9) and hypoxic training (Table 10). Athlete characteristics (Tables 11 and 12) produced significant changes, as well. Less-evident changes occurred due to pre-analytical conditions (Table 3) and acute exercise (Table 4).

**Table 1:**
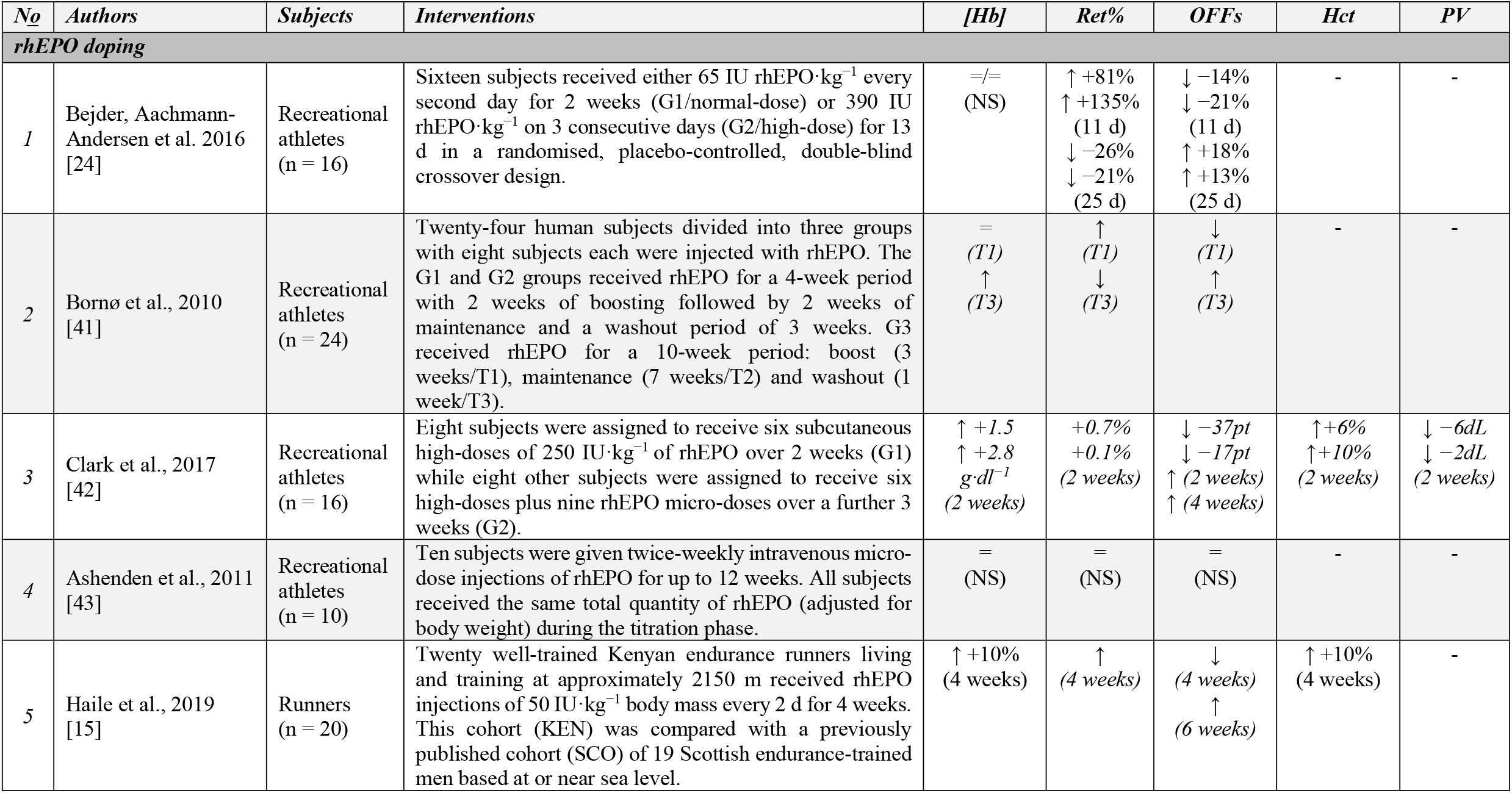
Changes of haematological variables related to rhEPO doping protocol. Numbers represent the relative changes during the most significant measurement: haemoglobin concentration ([Hb]), reticulocytes percentage (Ret%), OFF-Score (OFFS), haematocrit (Hct) and plasma volume (PV). Values in italics correspond to absolute variations.

**Table 2:**
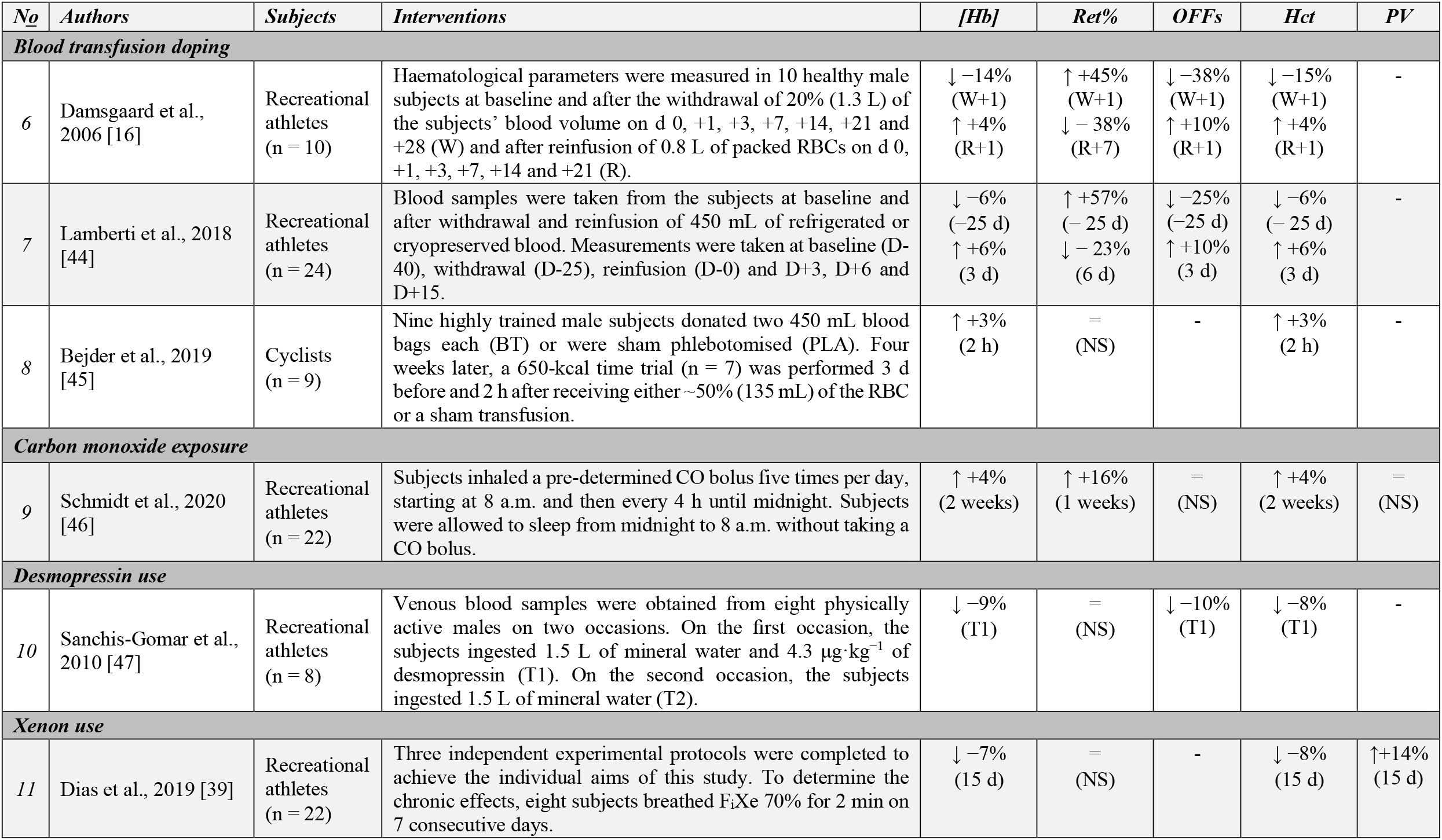
Changes of haematological variables related to various doping practises. Numbers represent the relative changes during the most significant measurement: haemoglobin concentration ([Hb]), reticulocytes percentage (Ret%), OFF-Score (OFFS), haematocrit (Hct) and plasma volume (PV).

| *No* | *Authors* | *Subjects* | *Interventions* | *[Hb]* | *Ret%* | *OFFs* | *Hct* | *PV* |
| --- | --- | --- | --- | --- | --- | --- | --- | --- |
| **Blood transfusion doping** |  |  |  |  |  |  |  |  |
| 6 | Damsgaard et al., 2006 [16] | Recreational athletes (n = 10) | Haematological parameters were measured in 10 healthy male subjects at baseline and after the withdrawal of 20% (1.3 L) of the subjects' blood volume on d 0, +1, +3, +7, +14, +21 and +28 (W) and after reinfusion of 0.8 L of packed RBCs on d 0, +1, +3, +7, +14 and +21 (R). | ↓ -14% (W+1)<br>↑ +4% (R+1) | ↑ +45% (W+1)<br>↓ - 38% (R+7) | ↓ -38% (W+1)<br>↑ +10% (R+1) | ↓ -15% (W+1)<br>↑ +4% (R+1) | - |
| 7 | Lamberti et al., 2018 [44] | Recreational athletes (n = 24) | Blood samples were taken from the subjects at baseline and after withdrawal and reinfusion of 450 mL of refrigerated or cryopreserved blood. Measurements were taken at baseline (D-40), withdrawal (D-25), reinfusion (D-0) and D+3, D+6 and D+15. | ↓ -6% (-25 d)<br>↑ +6% (3 d) | ↑ +57% (- 25 d)<br>↓ - 23% (6 d) | ↓ -25% (-25 d)<br>↑ +10% (3 d) | ↓ -6% (- 25 d)<br>↑ +6% (3 d) | - |
| 8 | Bejder et al., 2019 [45] | Cyclists (n = 9) | Nine highly trained male subjects donated two 450 mL blood bags each (BT) or were sham phlebotomised (PLA). Four weeks later, a 650-kcal time trial (n = 7) was performed 3 d before and 2 h after receiving either ~50% (135 mL) of the RBC or a sham transfusion. | ↑ +3% (2 h) | = (NS) | - | ↑ +3% (2 h) | - |
| **Carbon monoxide exposure** |  |  |  |  |  |  |  |  |
| 9 | Schmidt et al., 2020 [46] | Recreational athletes (n = 22) | Subjects inhaled a pre-determined CO bolus five times per day, starting at 8 a.m. and then every 4 h until midnight. Subjects were allowed to sleep from midnight to 8 a.m. without taking a CO bolus. | ↑ +4% (2 weeks) | ↑ +16% (1 weeks) | = (NS) | ↑ +4% (2 weeks) | = (NS) |
| **Desmopressin use** |  |  |  |  |  |  |  |  |
| 10 | Sanchis-Gomar et al., 2010 [47] | Recreational athletes (n = 8) | Venous blood samples were obtained from eight physically active males on two occasions. On the first occasion, the subjects ingested 1.5 L of mineral water and 4.3 µg·kg<sup>-1</sup> of desmopressin (T1). On the second occasion, the subjects ingested 1.5 L of mineral water (T2). | ↓ -9% (T1) | = (NS) | ↓ -10% (T1) | ↓ -8% (T1) | - |
| **Xenon use** |  |  |  |  |  |  |  |  |
| 11 | Dias et al., 2019 [39] | Recreational athletes (n = 22) | Three independent experimental protocols were completed to achieve the individual aims of this study. To determine the chronic effects, eight subjects breathed FiXe 70% for 2 min on 7 consecutive days. | ↓ -7% (15 d) | = (NS) | - | ↓ -8% (15 d) | ↑ +14% (15 d) |

**Table 3:** Changes of haematological variables related to pre-analytical variations. Numbers represent the relative changes during the most significant measurement: haemoglobin concentration ([Hb]), reticulocytes percentage (Ret%), OFF-Score (OFFS), haematocrit (Hct) and plasma volume (PV). Values in italics correspond to absolute variations.

| *No* | *Authors* | *Subjects* | *Interventions* | *[Hb]* | *Ret%* | *OFFs* | *Hct* | *PV* |
| --- | --- | --- | --- | --- | --- | --- | --- | --- |
| **Pre-analytical factors** |  |  |  |  |  |  |  |  |
| 12 | Robinson et al., 2019 [48] | Track and field athletes (n = 3683) | Reference ranges in top-level track and field athletes were determined for biomarkers of altered erythropoiesis considering various factors such as gender, age, endurance or non-endurance disciplines, detailed sport disciplines, the origin of the athletes (African vs non-African) and the time of blood sampling: morning (T1), afternoon (T2) and evening (T3). | ↓<br>(T2)<br>↓<br>(T3) | =<br>(NS) | ↓<br>(T2)<br>(T3) | - | - |
| 13 | Sennels et al., 2011 [25] | Recreational athletes (n = 24) | Venous blood samples were obtained under standardised circumstances from 24 healthy young men every third hour through 24 h for a total of nine samples. | ↑ +3%<br>(12:00) | ↑ +5%<br>(12:00)<br>*Ret count | - | ↑ +3%<br>(12:00) | - |
| 14 | Kuipers et al., 2005 [17] | Endurance +<br>Recreational athletes (n = 7) | In seven subjects in separate experiments, 500 mL of saline were infused around 8 a.m., and blood variables were measured before and every hour thereafter until 7 h after infusion. | ↓ -5%<br>(1 h) | =<br>(NS) | - | ↓ -4%<br>(1 h) | - |
| 15 | Bejder, Hoffmann et al., 2016 [31] | Recreational athletes (n = 20) | Twenty subjects received rhEPO for 3 weeks. After 10 d of rhEPO washout, 10 subjects ingested a 1000 mL bolus of water. Blood variables were measured +20, +40, +60 and +80 min after ingestion. | ↓ -3%<br>(60 min) | =<br>(NS) | ↑ -4%<br>(40 min) | - | ↑ +4%<br>(60 min) |
| 16 | Astolfi, Schumacher et al., 2020 [13] | Cyclists +<br>Divers +<br>Recreational athletes (n = 38) | Ten successive venous blood samples from 38 subjects were collected: after 10 min of normalised activity (B1), after 10 min seated (B2, typical reference sample in an anti-doping context), after a 50 m walk (B3), after an additional 5 and 10 min in a seated position (B4 and B5) and finally after 5–30 min in a supine position (B6-B10). | ↑ +3%<br>(B1) | =<br>(NS) | - | ↑ +4%<br>(B1) | ↓<br>(B1) |
| 17 | Schumacher et al., 2012 [49] | Endurance athletes (n = 15) | Fifteen endurance athletes submitted ABP blood samples in the morning before (T1) and after arrival (T2) on an 8 h flight. Two additional samples were obtained in the morning and the evening 3 d after travel. Twelve nontraveling subjects served as controls. | ↓ -0.5<br>g·dl<sup>-1</sup><br>(T2) | =<br>(NS) | - | ↓<br>(T2) | - |
| 18 | Alsaadi et al., 2015 [50] | Recreational athletes (n = 11) | Nine healthy, physically active subjects were tested twice daily—in the morning (T1) and afternoon (T2)—for 2 d before and 3 d into the celebration of Ramadan. Sample collection and all analyses were performed according to WADA technical documents. | ↓ -0.4<br>↓ -0.3<br>g·dl<sup>-1</sup><br>(T1/T2) | =<br>(NS) | - | - | - |

**Table 4:**
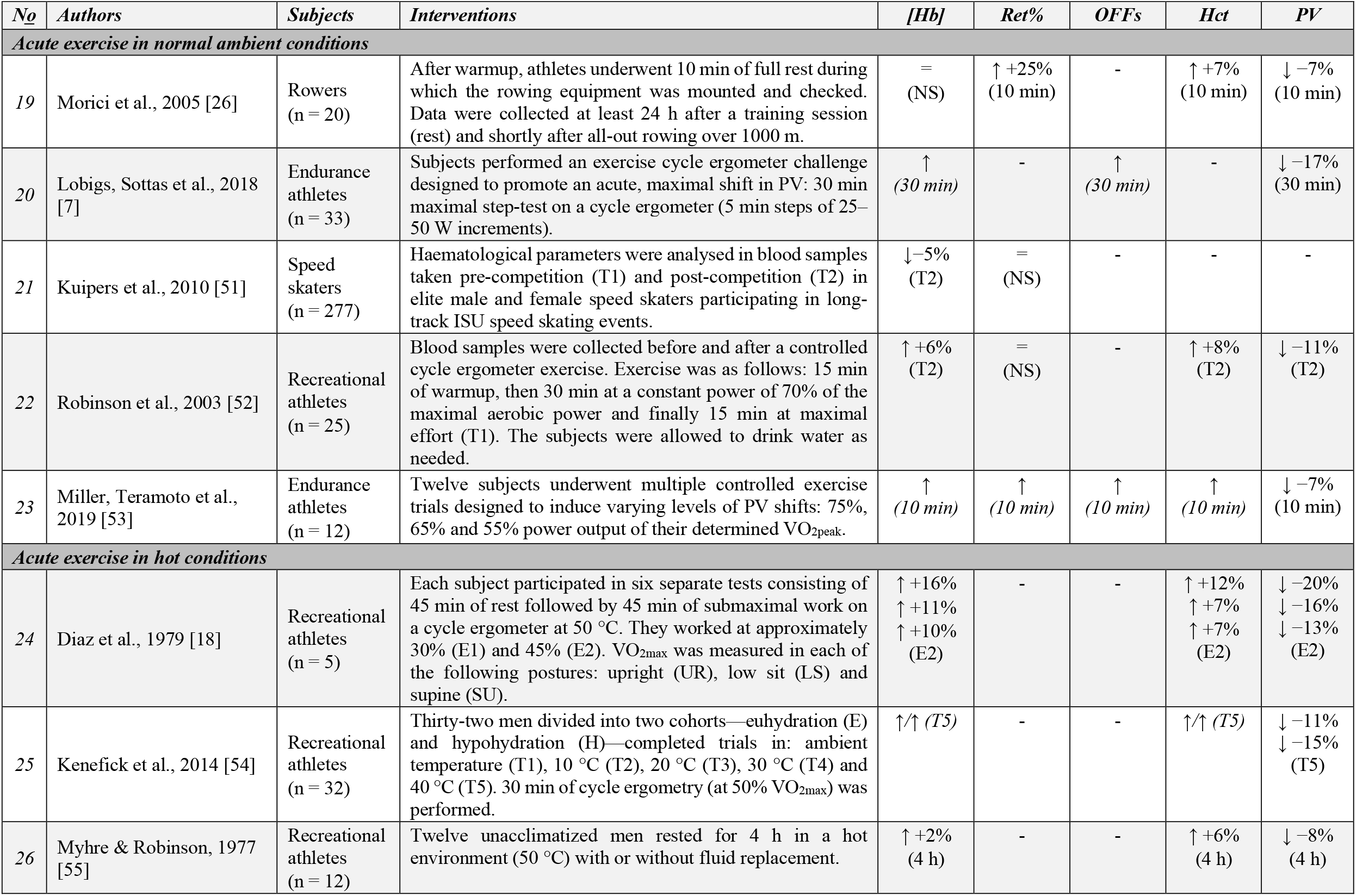
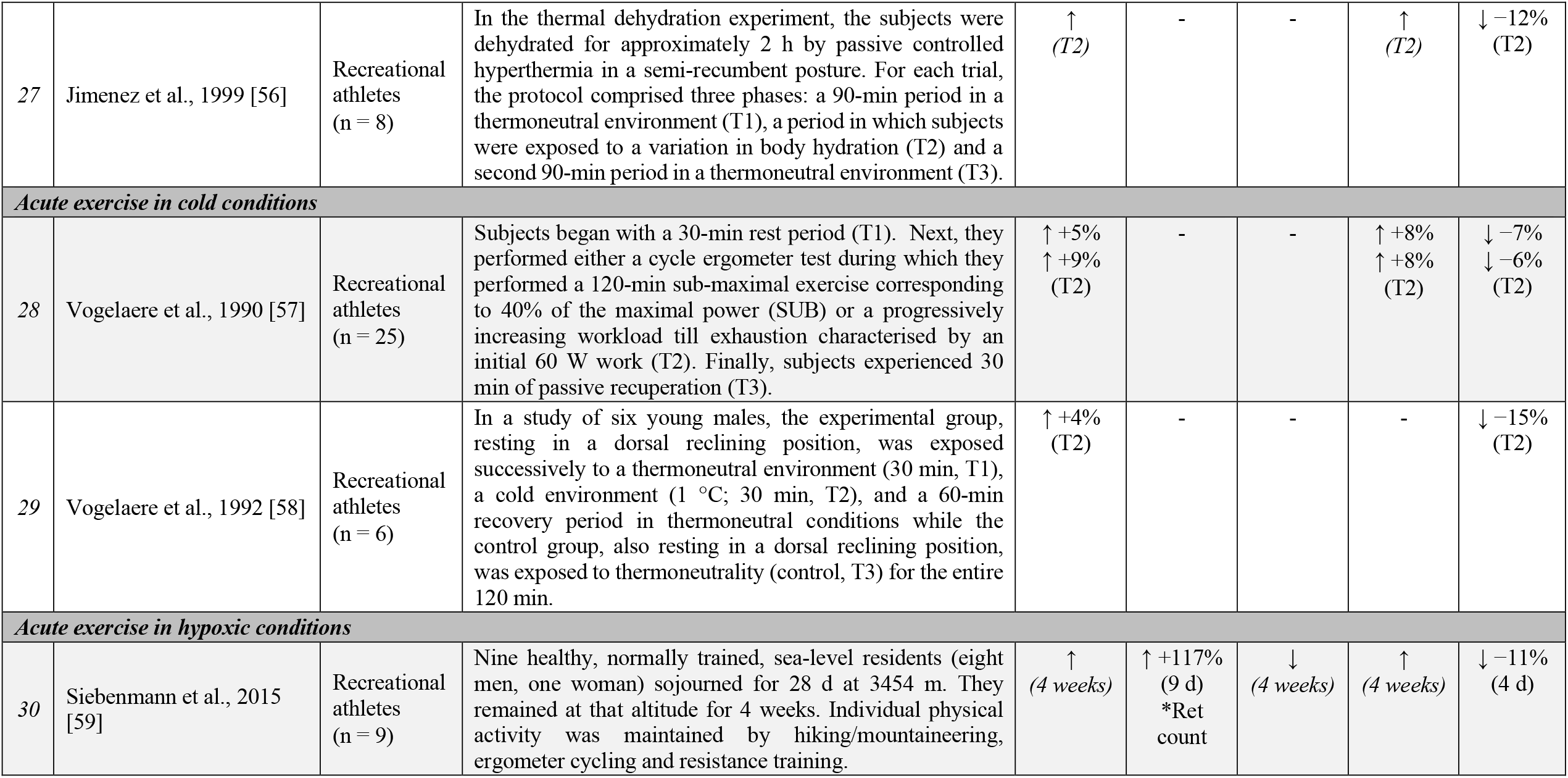
Changes of haematological variables related to acute exercises in various environmental conditions. Numbers represent the relative changes during the most significant measurement: haemoglobin concentration ([Hb]), reticulocytes percentage (Ret%), OFF-Score (OFFS), haematocrit (Hct) and plasma volume (PV). Values in italics correspond to absolute variations.

| No | Authors | Subjects | Interventions | [Hb] | Ret% | OFFs | Hct | PV |
| --- | --- | --- | --- | --- | --- | --- | --- | --- |
| **Acute exercise in normal ambient conditions** |  |  |  |  |  |  |  |  |
| 19 | Morici et al., 2005 [26] | Rowers (n = 20) | After warmup, athletes underwent 10 min of full rest during which the rowing equipment was mounted and checked. Data were collected at least 24 h after a training session (rest) and shortly after all-out rowing over 1000 m. | = (NS) | ↑ +25% (10 min) | - | ↑ +7% (10 min) | ↓ -7% (10 min) |
| 20 | Lobigs, Sottas et al., 2018 [7] | Endurance athletes (n = 33) | Subjects performed an exercise cycle ergometer challenge designed to promote an acute, maximal shift in PV: 30 min maximal step-test on a cycle ergometer (5 min steps of 25–50 W increments). | ↑ (30 min) | - | ↑ (30 min) | - | ↓ -17% (30 min) |
| 21 | Kuipers et al., 2010 [51] | Speed skaters (n = 277) | Haematological parameters were analysed in blood samples taken pre-competition (T1) and post-competition (T2) in elite male and female speed skaters participating in long-track ISU speed skating events. | ↓ -5% (T2) | = (NS) | - | - | - |
| 22 | Robinson et al., 2003 [52] | Recreational athletes (n = 25) | Blood samples were collected before and after a controlled cycle ergometer exercise. Exercise was as follows: 15 min of warmup, then 30 min at a constant power of 70% of the maximal aerobic power and finally 15 min at maximal effort (T1). The subjects were allowed to drink water as needed. | ↑ +6% (T2) | = (NS) | - | ↑ +8% (T2) | ↓ -11% (T2) |
| 23 | Miller, Teramoto et al., 2019 [53] | Endurance athletes (n = 12) | Twelve subjects underwent multiple controlled exercise trials designed to induce varying levels of PV shifts: 75%, 65% and 55% power output of their determined VO<sub>2peak</sub>. | ↑ (10 min) | ↑ (10 min) | ↑ (10 min) | ↑ (10 min) | ↓ -7% (10 min) |
| **Acute exercise in hot conditions** |  |  |  |  |  |  |  |  |
| 24 | Diaz et al., 1979 [18] | Recreational athletes (n = 5) | Each subject participated in six separate tests consisting of 45 min of rest followed by 45 min of submaximal work on a cycle ergometer at 50 °C. They worked at approximately 30% (E1) and 45% (E2). VO<sub>2max</sub> was measured in each of the following postures: upright (UR), low sit (LS) and supine (SU). | ↑ +16%<br>↑ +11%<br>↑ +10% (E2) | - | - | ↑ +12%<br>↑ +7%<br>↑ +7% (E2) | ↓ -20%<br>↓ -16%<br>↓ -13% (E2) |
| 25 | Kenefick et al., 2014 [54] | Recreational athletes (n = 32) | Thirty-two men divided into two cohorts—euhydration (E) and hypohydration (H)—completed trials in: ambient temperature (T1), 10 °C (T2), 20 °C (T3), 30 °C (T4) and 40 °C (T5). 30 min of cycle ergometry (at 50% VO<sub>2max</sub>) was performed. | ↑/↑ (T5) | - | - | ↑/↑ (T5) | ↓ -11%<br>↓ -15% (T5) |
| 26 | Myhre & Robinson, 1977 [55] | Recreational athletes (n = 12) | Twelve unacclimatized men rested for 4 h in a hot environment (50 °C) with or without fluid replacement. | ↑ +2% (4 h) | - | - | ↑ +6% (4 h) | ↓ -8% (4 h) |
| 27 | Jimenez et al., 1999 [56] | Recreational athletes<br>(n = 8) | In the thermal dehydration experiment, the subjects were dehydrated for approximately 2 h by passive controlled hyperthermia in a semi-recumbent posture. For each trial, the protocol comprised three phases: a 90-min period in a thermoneutral environment (T1), a period in which subjects were exposed to a variation in body hydration (T2) and a second 90-min period in a thermoneutral environment (T3). | ↑<br>(T2) | - | - | ↑<br>(T2) | ↓ -12%<br>(T2) |
| **Acute exercise in cold conditions** |  |  |  |  |  |  |  |  |
| 28 | Vogelaere et al., 1990 [57] | Recreational athletes<br>(n = 25) | Subjects began with a 30-min rest period (T1). Next, they performed either a cycle ergometer test during which they performed a 120-min sub-maximal exercise corresponding to 40% of the maximal power (SUB) or a progressively increasing workload till exhaustion characterised by an initial 60 W work (T2). Finally, subjects experienced 30 min of passive recuperation (T3). | ↑ +5%<br>↑ +9%<br>(T2) | - | - | ↑ +8%<br>↑ +8%<br>(T2) | ↓ -7%<br>↓ -6%<br>(T2) |
| 29 | Vogelaere et al., 1992 [58] | Recreational athletes<br>(n = 6) | In a study of six young males, the experimental group, resting in a dorsal reclining position, was exposed successively to a thermoneutral environment (30 min, T1), a cold environment (1 °C; 30 min, T2), and a 60-min recovery period in thermoneutral conditions while the control group, also resting in a dorsal reclining position, was exposed to thermoneutrality (control, T3) for the entire 120 min. | ↑ +4%<br>(T2) | - | - | - | ↓ -15%<br>(T2) |
| **Acute exercise in hypoxic conditions** |  |  |  |  |  |  |  |  |
| 30 | Siebenmann et al., 2015 [59] | Recreational athletes<br>(n = 9) | Nine healthy, normally trained, sea-level residents (eight men, one woman) sojourned for 28 d at 3454 m. They remained at that altitude for 4 weeks. Individual physical activity was maintained by hiking/mountaineering, ergometer cycling and resistance training. | ↑<br>(4 weeks) | ↑ +117%<br>(9 d)<br>*Ret count | ↓<br>(4 weeks) | ↑<br>(4 weeks) | ↓ -11%<br>(4 d) |

**Table 5:** Changes of haematological variables related to prolonged and multiday events in ambient conditions. Numbers represent the relative changes during the most significant measurement: haemoglobin concentration ([Hb]), reticulocytes percentage (Ret%), OFF-Score (OFFS), haematocrit (Hct) and plasma volume (PV). Values in italics correspond to absolute variations.

| *No* | *Authors* | *Subjects* | *Interventions* | *[Hb]* | *Ret%* | *OFFs* | *Hct* | *PV* |
| --- | --- | --- | --- | --- | --- | --- | --- | --- |
| ***Prolonged exercise in normal ambient conditions*** |  |  |  |  |  |  |  |  |
| 31 | Miller, Beharry et al., 2019 [60] | Triathletes (n = 19) | Complete blood count tests were conducted on 19 Ironman triathletes before and after an Ironman triathlon to characterise changes in haematological parameters. | ↓<br>(2 d) | ↓<br>(2 d) | ↓<br>(2 d) | ↓<br>(2 d) | ↑+16%<br>(2 d) |
| 32 | Schumacher et al., 2003 [32] | Cyclists (n = 39) | Twenty-three male professional cyclists and 16 inactive control subjects were investigated during a 5 d road cycling stage race at sea level. Samples were obtained every morning prior to the race and between +1 and +3 h after the end of each stage. | ↓ -6%<br>(5 d) | =<br>(NS) | ↓ -15%<br>(4 d) | ↓ -5%<br>(4 d) | ↑ +5%<br>(5 d) |
| 33 | Fallon et al., 1999 [27] | Runners (n = 9) | Blood samples were obtained from seven male and two female participants in a 1600 km ultramarathon foot race before (T1), after +4 d (T2) and +11 d (T3) of running and at the end of the race (T4). | ↓ -10%<br>(T3) | ↑ +63%<br>(T3) | - | - | ↑ +17%<br>(T3) |
| 34 | Mørkeberg et al., 2009 [61] | Cyclists (n = 28) | From December 2006 to November 2007, 374 blood samples and 287 urine samples were obtained from 28 elite male cyclists from the Danish cycling team, Team CSC. Seven riders were measured once before the race and twice during the Tour de France. | ↓ -12%<br>(19 d) | =<br>(NS) | ↓<br>(19 d) | ↓<br>(19 d) | ↑<br>(19 d) |
| 35 | Voss et al., 2013 [19] | Cyclists (n = 12) | Twelve highly trained cyclists tapered for 3 d before 6 d of simulated intense stage racing. Samples were taken in the morning and afternoon. | ↓ -13%<br>(8 d) | =<br>(NS) | - | ↓<br>(8 d) | ↑ +24%<br>(8 d) |
| 36 | Corsetti et al., 2012 [62] | Cyclists (n = 9) | Nine professional cyclists engaged in the 2011 Giro d'Italia stage race. Haematological parameters were measured d -1 (pre-race), d +12 and d +22 during the race. | ↓ -9%<br>(12 d) | =<br>(NS) | - | ↓ -7%<br>(12 d) | ↑ +2%<br>(12 d) |
| 37 | Lombardi, Lanteri, Fiorella et al., 2013 [63] | Cyclists (n = 253) | The study population was comprised of male professional cyclists competing in the 2010 (n = 144) and 2012 (n = 109) GiroBio 10-d stage races. Blood samples were taken before the start of the race (T1), at mid-race (T2) and at the end of the race (T3). Results are the average of the two races. | ↓ -9%<br>(T2) | ↓ -6%<br>(T2)<br>↑ +15%<br>(T3) | ↓↑<br>(T2)<br>(T3) | ↓↑<br>(T2)<br>(T3) | - |

**Table 6:** Changes of haematological variables related to prolonged and multiday events in specific conditions. Numbers represent the relative changes during the most significant measurement: haemoglobin concentration ([Hb]), reticulocytes percentage (Ret%), OFF-Score (OFFS), haematocrit (Hct) and plasma volume (PV). Values in italics correspond to absolute variations.

| No | Authors | Subjects | Interventions | [Hb] | Ret% | OFFs | Hct | PV |
| --- | --- | --- | --- | --- | --- | --- | --- | --- |
| **Prolonged exercise in hot conditions** |  |  |  |  |  |  |  |  |
| 38 | Rama et al., 2015 [36] | Runners<br>(n = 19) | The race was conducted over five stages (5 d) totalling a distance of 230 km. The daily maximum temperature ranged between 32 and 40 °C. Pre-stage blood samples were collected within the hour prior to the start of each running stage. | ↓ -7%<br>(5 d) | - | - | ↓ -8%<br>(5 d) | ↑ +18%<br>(5 d) |
| 39 | Zappe et al., 1993 [64] | Cyclists<br>(n = 9) | Nine subjects were assigned to one of two experimental treatments: euhydrated (E) and hypohydrated (H). Following 20 min of a seated test in a warm environment, each subject cycled in a semi-reclining posture for 60 min at three successive intensities, representing 22% (T1), 37% (T2) and 53% (T3) of the VO<sub>2max</sub>. | ↑/↑<br>(T3) | - | - | ↑/↑<br>(T3) | ↓ -11%<br>↓ -11%<br>(T3) |
| 40 | Gaebelein & Senay, 1980 [65] | Recreational athletes<br>(n = 4) | Four males were studied during cycle ergometer exercise and stair stepping in a hot, wet environment (32 °C) after exertion. Venous blood samples were obtained 24 h before each exercise and before and at 10 min intervals during each exercise. | ↑<br>(10 min) | - | - | ↑<br>(10 min) | ↓<br>(10 min) |
| **Prolonged exercise in hypoxic conditions** |  |  |  |  |  |  |  |  |
| 41 | Garvican-Lewis et al., 2014 [66] | Cyclists<br>(n = 30) | Four teams participating in the 2013 Tour of Qinghai Lake agreed to participate in the study. Haematological variables of sea-level (SL) and altitude (ALT) cyclists were measured during a 14-d cycling race, held at 1146–4120 m, as well as during a simulated tour near sea level (SIM). | ↓ -4%<br>(10 d)<br>↓ -8%<br>↓ -7%<br>(14 d) | ↑ +28%<br>↑ +24%<br>(10 d)<br>↓ -18%<br>↓ -35%<br>(14 d) | - | ↓ -7%<br>(6 d)<br>↓ -6%<br>↓ -7%<br>(14 d) | ↑ +15%<br>(6 d)<br>↑ +17%<br>↑ +16%<br>(14 d) |
| 42 | Schumacher et al., 2015 [67] | Cyclists<br>(n = 25) | Fourteen sea-level (SL) and eleven altitude-native (ALT), highly trained athletes participated in a 14-d cycling stage race taking place at an average altitude of 2496 m above sea level (min. 1014 m, max. 4120 m). Race distances ranged between 96 and 227 km d<sup>-1</sup>. Blood samples were taken on d -1, +3, +6, +10, +14 (SL) and -1, +9, +15 (ALT). | ↓<br>(14 d) | ↑<br>(14 d) | ↓<br>(14 d) | - | ↑<br>(14 d) |

**Table 7:** Changes of haematological variables related to various forms of chronic training. Numbers represent the relative changes during the most significant measurement: haemoglobin concentration ([Hb]), reticulocytes percentage (Ret%), OFF-Score (OFFS), haematocrit (Hct) and plasma volume (PV). Values in italics correspond to absolute variations.

| *No* | *Authors* | *Subjects* | *Interventions* | *[Hb]* | *Ret%* | *OFFs* | *Hct* | *PV* |
| --- | --- | --- | --- | --- | --- | --- | --- | --- |
| **Endurance training** |  |  |  |  |  |  |  |  |
| 43 | Spodaryk, 1993 [68] | Various sports (n = 39) | Haematological and iron-related parameters were measured at rest in 39 male athletes from the Polish team who participated in the 1988 Seoul Olympics. The athletes were divided into two groups: endurance-trained subjects (EN: cyclists, canoeists and rowers) and strength-trained subjects (ST: wrestlers and judoka). | Lower (EN) | Higher (EN) | - | - | - |
| 44 | Bejder et al., 2017 [33] | Cyclists (n = 11) | Eleven high-level competitive cyclists were investigated for 3 weeks. After initial measurements in week 1 (T1), training load was increased ~250% in week 2 (T2) followed by a reversion to the baseline training load in week 3 (T3). PV and haematological variables were determined frequently during all weeks. | ↓ -6% (T2) | ↑ +15% (T2) | ↓ -16% (T2) | ↓ -5% (T2) | ↑ +10% (T2) |
| 45 | Green et al., 1991 [69] | Recreational athletes (n = 8) | Eight healthy, active but untrained males volunteered for the training study. The training program involved 8 weeks of cycle exercise, with each training session lasting for 2 h and with the intensity initially set at 62% of each subject's VO<sub>2max</sub>. Training was performed on a 3-l-3 cycle, with exercise conducted for 3 consecutive days followed by 1 d of inactivity and another 3 consecutive days of exercise. | ↓ -4% (4 weeks) | = (NS) | - | ↓ -5% (4 weeks) | ↑ +14% (4 weeks) |
| 46 | Montero et al., 2017 [20] | Untrained athletes (n = 9) | Nine healthy, untrained volunteers underwent supervised endurance training consisting of 3–4 × 60 min cycle ergometry sessions per week for 8 weeks. Haematological markers were determined before and at weeks +2 (T1), +4 (T2) and +8 (T3) of training. | ↓ -8% (T1) | = (NS) | - | ↓ -7% (T1) | ↑ +16% (T1) |
| **Apnea training** |  |  |  |  |  |  |  |  |
| 47 | Revelli et al., 2013 [28] | Divers (n = 6) | Six scuba divers were tested before, during and after a 14-d Guinness saturation dive (8–10 m). During the dive, athletes breathed air at 1.8–2 ATA under the control of a team of physicians. Serum parameters were measured before (T0), during (T1, T2) and after the dive (T3, T4). | = (NS) | ↓ -44% (T3) | - | ↓ -6% (T2) | - |

**Table 8:** Changes in haematological variables related to training periodisation. Numbers represent the relative changes during the most significant measurement: haemoglobin concentration ([Hb]), reticulocytes percentage (Ret%), OFF-Score (OFFS), haematocrit (Hct) and plasma volume (PV). Values in italics correspond to absolute variations.

| No | Authors | Subjects | Interventions | [Hb] | Ret% | OFFs | Hct | PV |
| --- | --- | --- | --- | --- | --- | --- | --- | --- |
| **Training content** |  |  |  |  |  |  |  |  |
| 48 | Banfi & Del Fabbro, 2007 [70] | Various sports (n = 63) | The behaviour of reticulocyte and immature reticulocyte fraction (IRF) were analysed in top-level athletes practising rugby (RUG), ski (SKI), soccer (SOC) and cycling (CYC) throughout a competitive season. Data were collected three to four times: before the start of the training period (T1), at the beginning (T2), in the middle (T3) and at the end of the competitive season (T4). | Lower<br>(RUG) | Lower<br>(RUG)<br>(CYC)<br>(SOC)<br>Higher<br>(SKI) | - | - | - |
| 49 | Abellan, Ventura, Pichini et al., 2007 [71] | Various sports (n = 246) | Haematological parameters were measured in 96 elite athletes of various sports. Elite athletes participated in different sports (swimming, synchronised swimming, taekwondo, rhythmic gymnastics, soccer, triathlon and weightlifting). | =<br>(NS) | =<br>(NS) | =<br>(NS) | =<br>(NS) | =<br>(NS) |
| 50 | Malcovati et al., 2003 [72] | Football players (n = 923) | This study tested the effect of age, ethnicity, exercise modalities and training phases on haematologic parameters and then estimated components of variation. Differences between low- (T1), int- (T2) and high-season (T2) were analysed. | ↓<br>(T2) | ↓<br>(T2) | - | ↓<br>(T2) | - |
| 51 | Banfi et al., 2006 [73] | Rugby players (n = 19) | Blood samples were collected at four consecutive training camps during a whole competitive season—first, at the start of the training period (T1), then after the training meeting (T2), after the first part of the championships (T3) and finally, at the end of the championships (T4). | ↓ -3%<br>(T3) | ↓ -21%<br>(T4) | - | ↓ -5%<br>(T3) | - |
| 52 | Banfi et al., 2010 [74] | Skiers (n = 18) | Haematological variations of elite skiers were monitored over four seasons. The study sample included 18 alpine skiers of the Italian National Alpine Ski Team (10 men and eight women). Skiers began training in July (T1) and competed from November till April (T2). | ↓<br>(T2) | ↑<br>(T1) | - | - | - |
| **Taper period** |  |  |  |  |  |  |  |  |
| 53 | Mujika et al., 2000 [75] | Runners (n = 8) | After 15 weeks of training, athletes completed a low-volume taper, consisting of either a 75% progressive reduction in pre-taper, low-intensity continuous training or high-intensity interval training. Blood samples were obtained before (T1) and after (T2) the taper period. | ↓ -3%<br>(T2) | ↑ +44%<br>(T2) | - | =<br>(NS) | - |
| 54 | Mujika et al., 2002 [76] | Runners (n = 10) | After 18 weeks of training, nine male runners were assigned to a high-frequency taper (HFT) or a moderate frequency taper (MFT), consisting of training daily or resting every third day of the taper. Blood samples were obtained before (T1) and after (T2) the taper period. | =<br>(NS) | =<br>(NS) | - | =<br>(NS) | - |

**Table 9:** Changes of haematological variables related to thermal acclimation protocols. Numbers represent the relative changes during the most significant measurement: haemoglobin concentration ([Hb]), reticulocytes percentage (Ret%), OFF-Score (OFFS), haematocrit (Hct) and plasma volume (PV). Values in italics correspond to absolute variations.

| No | Authors | Subjects | Interventions | [Hb] | Ret% | OFFs | Hct | PV |
| --- | --- | --- | --- | --- | --- | --- | --- | --- |
| **Heat and cold acclimation/acclimatisation** |  |  |  |  |  |  |  |  |
| 55 | Costa et al., 2014 [77] | Runners (n = 6) | Three running exercise-heat exposures were performed at 30°C (T1), followed by three running exercise-heat exposures at 35 °C (T2). Each running exercise-heat exposure was separated by 48 h. | ↓<br>(T1) | - | - | ↓<br>(T1) | ↑ +7%<br>(T1) |
| 56 | Loeppky, 2015 [78] | Recreational athletes (n = 18) | PV variations were reported after 10 d of heat acclimation from studies in winter (WIN, n = 8) and summer (SUM, n = 10). In both studies, heat acclimation and dehydrating ergometer exercise were performed at 50 °C dry bulb and 26 °C wet bulb at 30% of each subject's VO<sub>2max</sub>. | =<br>↓ -2%<br>(10 d) | - | - | =/= (10 d) | =<br>↑ +5%<br>(10 d) |
| 57 | Scoon et al., 2007 [37] | Runners (n = 6) | Six male distance runners completed 3 weeks of post-training sauna bathing and 3 weeks of control training with a 3-week washout. During the sauna period, subjects sat in a humid sauna at 90 °C immediately post-exercise for 30 min on 12 occasions. | ↓ -1.3%<br>(3 weeks) | - | - | ↓ -2%<br>(3 weeks) | ↑ +7%<br>(3 weeks) |
| 58 | Stanley et al., 2015 [40] | Cyclists (n = 7) | Seven well-trained male cyclists were monitored for 35 consecutive days (17 d baseline training, 10 d training plus sauna and 8 d training). Sauna exposure consisted of 30 min (87 °C, 11% relative humidity) immediately following normal training. | ↓<br>(4 d) | - | - | ↓<br>(4 d) | ↑ +18%<br>(4 d) |
| 59 | Tebeck et al., 2020 [79] | Cyclists (n = 11) | Eleven cyclists completed each of two 5-d blocks of short-term heat acclimation matched for heat index (44 °C) and total exposure time (480 min). The blocks were separated by 30 d. Two protocols were applied: dry (D/43 °C and 20% humidity) and humid (H/32 °C and 80% humidity). | ↓/↓<br>(5 d) | - | - | ↓/↓<br>(5 d) | ↑ +5%<br>↑ +5%<br>(5 d) |
| 60 | Oberholzer et al., 2019 [80] | Cyclists (n = 21) | Participants completed the 5½-week heat acclimation period, after which blood sampling was repeated. Blood sampling was conducted after 2 weeks into the intervention period prior to an exercise training session. | =<br>(NS) | =<br>(NS)<br>*Ret count | - | =<br>(NS) | ↑ +7%<br>(2 weeks) |
| 61 | Banfi et al., 2008 [81] | Rugby players (n = 10) | Ten athletes were exposed daily to whole-body cryotherapy for 5 d at the Center of Spała, Poland. During this period, moderate training (3 h daily) continued for all athletes. | ↓ -2%<br>(5 d) | =<br>(NS) | - | =<br>(NS) | - |
| 62 | Lombardi, Lanteri, Porcelli et al., 2013 [34] | Rugby players (n = 27) | Twenty-seven professional rugby players received two daily cryostimulation treatments for 7 consecutive days (-110 °C to -140 °C). Blood samples were collected before and after the cryotherapeutic protocol. | ↓ -3%<br>(7 d) | =<br>(NS) | ↓ -5%<br>(7 d) | ↓ -1%<br>(7 d) | =<br>(NS) |
| 63 | Checinska-Maciejewska et al., 2019 [21] | Swimmers (n = 34) | Thirty-four healthy subjects (18 men and 16 women) aged 50 years swam in cold seawater during the winter season at least twice a week. The average water temperature was 10 °C in October (T1), 1 °C in January (T2) and 4 °C at the end of April (T3). | ↑ +9%<br>(T3) | - | - | =<br>(NS) | - |

**Table 10:** Changes of haematological variables related to hypoxic training strategies. Numbers represent the relative changes during the most significant measurement: haemoglobin concentration ([Hb]), reticulocytes percentage (Ret%), OFF-Score (OFFS), haematocrit (Hct) and plasma volume (PV). Values in italics correspond to absolute variations.

| No | Authors | Subjects | Interventions | [Hb] | Ret% | OFFs | Hct | PV |
| --- | --- | --- | --- | --- | --- | --- | --- | --- |
| **Living high-training high (LHTH) protocols** |  |  |  |  |  |  |  |  |
| 64 | Bonne et al., 2015 [35] | Swimmers (n = 20) | One group of swimmers lived high and trained high (LHTH, n = 10) for three to four weeks at 2130 m or higher, while a control group (n = 10) completed a three-week training camp at sea level. Haematological parameters were determined weekly three times before and four times after the training camps (+7, +14, +21 and +28 d). | =<br>(NS) | ↓ -34%<br>(7 d) | ↑ +22%<br>(7 d) | ↑ +5%<br>(7 d) | - |
| 65 | Garvican et al., 2012 [82] | Cyclists (n = 13) | Haematological parameters were measured in 13 elite cyclists during and 10 d after 3 weeks of sea level or altitude training (2760 m). The altitude group participated in a 3-week natural altitude training camp at Passo dello Stelvio, Italy, living at 2760 m and training during the majority of ride time > 1800 m for 2–6 h d<sup>-1</sup>. | =<br>(NS) | ↑ +25%<br>↑ (12 d)<br>↓ (31 d) | - | =<br>(NS) | - |
| 66 | Wachsmuth et al., 2013 [22] | Swimmers (n = 27) | Twenty-five athletes trained between one and three times for 3–4 weeks at altitude training camps. Three training camp were at 2320 m (G1) in Sierra Nevada, Spain, and one training camp was at 1360 m (G2) in Pretoria, South Africa. | =<br>↑ +4%<br>(4 weeks) | =/= (NS) | =/= (NS) | =/= (NS) | =/= (NS) |
| 67 | Ashenden et al., 2001 [83] | Endurance athletes (n = 23) | The haematological and physiological responses of 23 well-trained athletes (12 cyclists, 10 kayakers, 13 triathletes and 11 middle-distance runners) exposed to a simulated altitude of 2650 m for 11 ± 23 nights (ALT) were contrasted with those of healthy volunteers receiving a low dose (150 IU·kg<sup>-1</sup> per week) of rEPO for 25 d (DOP). | - | ↑ +36%<br>(17 d) | - | - | - |
| **Living high-training low (LHTL) protocols** |  |  |  |  |  |  |  |  |
| 68 | Ashenden et al., 2003 [84] | Endurance athletes (n = 64) | Haematologic data were collected from three groups: 19 elite cyclists who lived and trained 2690 m above sea level for 26–31 d (LH-TH), from 39 well-trained subjects who resided at sea level but slept at a simulated altitude of 2650–3000 m for 20–23 d of consecutive or intermittent nightly exposure (LH-TL), and from six elite Kenyan runners who lived 2100 m above sea level but descended to compete at sea level competitions. | ↑<br>(21 d) | ↑<br>(21 d) | ↑<br>(21 d) | - | - |
| 69 | Garvican-Lewis et al., 2018 [85] | Endurance athletes (n = 34) | Thirty-four endurance-trained athletes (20 runners, 11 cyclists and three triathletes) completed 3 weeks of simulated LH-TL altitude training, accumulating an average of 14 h d<sup>-1</sup> at a simulated altitude (normobaric hypoxia) of 3000 m. Athletes received either oral (ORAL), intravenous (IV) or placebo iron supplementation (PLA). | ↑ +0,9<br>↑ +0.8<br>↑ +0.6<br>g·dl<sup>-1</sup><br>(21 d) | ↓ -0.2%<br>↓ -0.2%<br>= (21 d) | ↑ +10 pt<br>↑ +8 pt<br>↑ +9 pt<br>(21 d) | - | - |
| 70 | Neya et al., 2013 [86] | Recreational athletes (n = 14) | Fourteen male collegiate runners were equally divided into two groups: altitude (ALT) and control (CON). Both groups spent 22 d at 1300–1800 m. The ALT group spent 10 h/night for 21 nights in simulated altitude (3000 m), while the CON group stayed at 1300 m (Takayama, Gifu, Japan). | ↑<br>(17 d) | ↑<br>(17 d) | - | ↑<br>(17 d) | - |
| 71 | Voss et al., 2020 [87] | Runners (n = 10) | The participants spent $\sim 6 \text{ h} \cdot \text{d}^{-1}$ at 3000–5400 m during waking hours and $\sim 10 \text{ h} \cdot \text{d}^{-1}$ overnight at 2400–3000 m simulated altitude. Venous blood samples were collected before hypoxic exposure (B0), after +1 (D1), +4 (D4), +7 (D7) and +14 (D14) d of hypoxic exposure and again +14 d post-exposure (P14). | $\uparrow +0,9 \text{ g} \cdot \text{dl}^{-1}$ (D14)<br>$\downarrow -1,3 \text{ g} \cdot \text{dl}^{-1}$ (P14) | $\uparrow +0.4\%$ (D7)<br>↓ (P14) | ↑ (P14) | - | - |
| **Intermittent hypoxic exposure (IHE) or training (IHT)** |  |  |  |  |  |  |  |  |
| 72 | Kasperska & Zembron-Lacny, 2020 [29] | Wrestlers (n = 12) | Twelve wrestlers were assigned to two groups: hypoxia (sports training combined with intermittent hypoxic exposure) and control (sports training only). An approximately 1 h intermittent hypoxic exposure was performed once a day for 10 d with one day off after 6 d. | = (NS) | $\uparrow +350\%$ (10 d) | - | = (NS) | - |
| 73 | Abellan, Ventura, Remacha et al., 2007 [88] | Triathletes (n = 16) | Sixteen male triathletes were randomly assigned to either the intermittent hypoxia exposure group or the control normoxic group. The exposure group were exposed to simulated altitude (4000–5500 m) in a hypobaric chamber for 3 h $\text{d}^{-1}$, 5 d a week for 4 weeks. Blood and urine samples were collected before and after the first (T0) and the final exposures (T1) and 2 weeks after the final exposure (T2). | = (NS) | = (NS) | - | = (NS) | - |
| 74 | Sanchez & Borrani, 2018 [89] | Runners (n = 30) | Fifteen highly trained endurance runners completed a 6-week regimented training with three sessions per week consisting of intermittent runs ($6 \times$ work-rest ratio of 5':5') on a treadmill at 80–85% of maximal aerobic speed. Nine athletes (the hypoxic group) performed the exercise bouts at $\text{FI}0_2 = 10.6\text{--}11.4\%$, while six athletes (the normoxic group) exercised in ambient air. | = (NS) | = (NS) | = (NS) | = (NS) | - |

**Table 11:** Changes of haematological variables related to athletes’ disorders or diseases. Numbers represent the relative changes during the most significant measurement: haemoglobin concentration ([Hb]), reticulocytes percentage (Ret%), OFF-Score (OFFS), haematocrit (Hct) and plasma volume (PV). Values in italics correspond to absolute variations.

| *No* | *Authors* | *Subjects* | *Interventions* | *[Hb]* | *Ret%* | *OFFs* | *Hct* | *PV* |
| --- | --- | --- | --- | --- | --- | --- | --- | --- |
| **Haematological disorders** |  |  |  |  |  |  |  |  |
| 75 | Parisotto et al., 2003 [90] | Elite athletes (n = 1152) | Blood samples were obtained from elite athletes from 12 countries with known haematologic abnormalities (G1). The algorithm scores for these athletes were compared with those of their healthy counterparts (G2) in terms of anaemia, iron deficiency and haemoglobinopathies. | Lower (G1) | Higher (G1) | Lower (G1) | Lower (G1) | - |
| 76 | Stangerup et al., 2017 [30] | Recreational athletes (n = 18) | Blood variables were measured at baseline and +3, +7, +14, +21 and +28 d after blood donation in 18 iron-sufficient women. | ↓ -8% (3 d) | ↑ +27% (7 d)<br>*Ret count | - | ↓ -8% (3 d) | - |
| 77 | Meurrens, 2016 [23] | Recreational athletes (n = 24) | Twenty-four moderately trained subjects were randomly divided into a donation (n = 16) and a placebo (n = 8) group. The three donations were spaced 3 months apart, and the recovery of endurance capacity and haematological parameters was monitored up to 1 month after the donation. | ↓ -12% (2 d) | - | - | ↓ -11% (2 d) | - |
| 78 | Ryan et al., 2016 [38] | Recreational athletes (n = 7) | Haematological values were measured for seven healthy, recreationally active men before (T1), 5 h after (T2) and 5 d (T3) after 4 d of head-down tilt bed rest. | ↑ +11% (T2) | = (NS)<br>*Ret count | - | ↑ +11% (T2) | ↓ -14% (T2) |

**Table 12:**
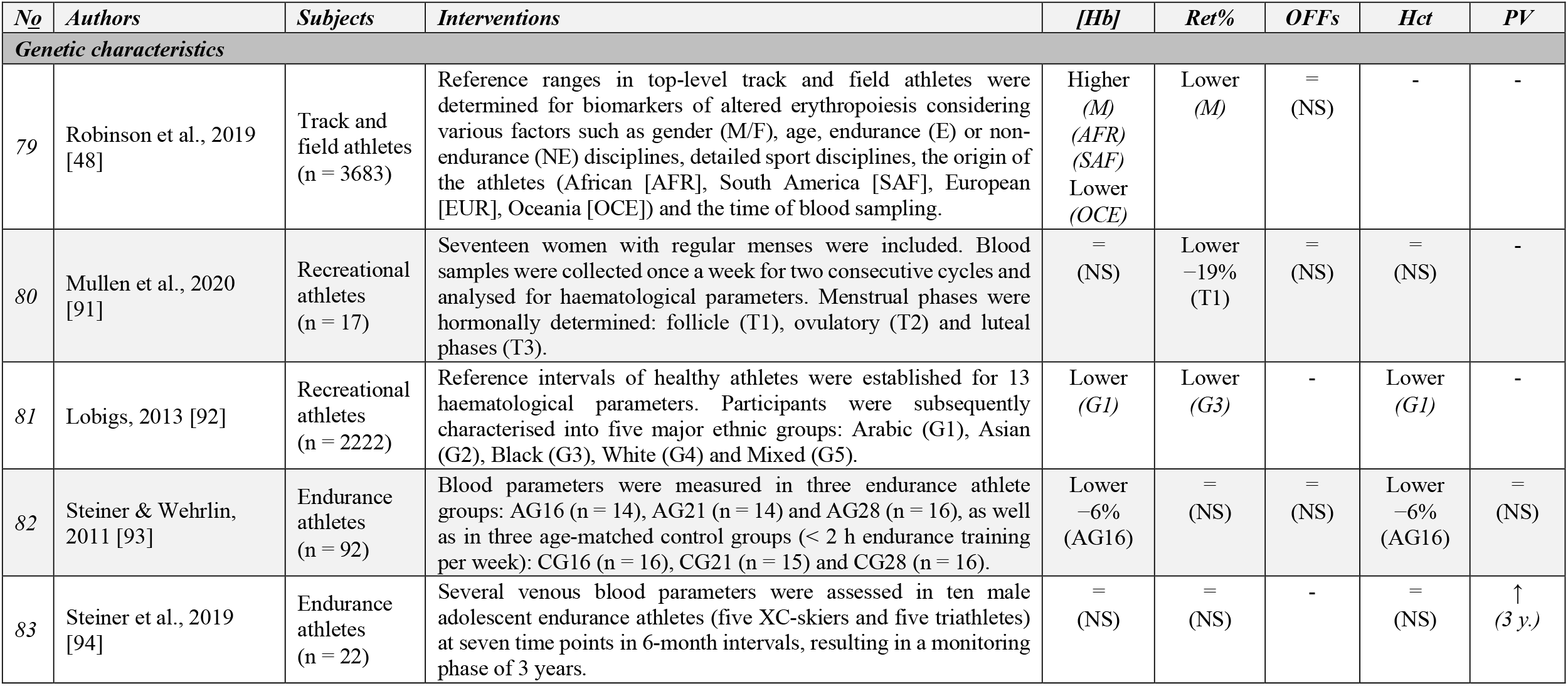
Changes in haematological variables related to athletes’ characteristics. Numbers represent the relative changes during the most significant measurement: haemoglobin concentration ([Hb]), reticulocytes percentage (Ret%), OFF-Score (OFFS), haematocrit (Hct) and plasma volume (PV). Values in italics correspond to absolute variations.

| No | Authors | Subjects | Interventions | [Hb] | Ret% | OFFs | Hct | PV |
| --- | --- | --- | --- | --- | --- | --- | --- | --- |
| **Genetic characteristics** |  |  |  |  |  |  |  |  |
| 79 | Robinson et al., 2019 [48] | Track and field athletes (n = 3683) | Reference ranges in top-level track and field athletes were determined for biomarkers of altered erythropoiesis considering various factors such as gender (M/F), age, endurance (E) or non-endurance (NE) disciplines, detailed sport disciplines, the origin of the athletes (African [AFR], South America [SAF], European [EUR], Oceania [OCE]) and the time of blood sampling. | Higher<br>(M)<br>(AFR)<br>(SAF)<br>Lower<br>(OCE) | Lower<br>(M) | =<br>(NS) | - | - |
| 80 | Mullen et al., 2020 [91] | Recreational athletes (n = 17) | Seventeen women with regular menses were included. Blood samples were collected once a week for two consecutive cycles and analysed for haematological parameters. Menstrual phases were hormonally determined: follicle (T1), ovulatory (T2) and luteal phases (T3). | =<br>(NS) | Lower<br>-19%<br>(T1) | =<br>(NS) | =<br>(NS) | - |
| 81 | Lobigs, 2013 [92] | Recreational athletes (n = 2222) | Reference intervals of healthy athletes were established for 13 haematological parameters. Participants were subsequently characterised into five major ethnic groups: Arabic (G1), Asian (G2), Black (G3), White (G4) and Mixed (G5). | Lower<br>(G1) | Lower<br>(G3) | - | Lower<br>(G1) | - |
| 82 | Steiner & Wehrlin, 2011 [93] | Endurance athletes (n = 92) | Blood parameters were measured in three endurance athlete groups: AG16 (n = 14), AG21 (n = 14) and AG28 (n = 16), as well as in three age-matched control groups (< 2 h endurance training per week): CG16 (n = 16), CG21 (n = 15) and CG28 (n = 16). | Lower<br>-6%<br>(AG16) | =<br>(NS) | =<br>(NS) | Lower<br>-6%<br>(AG16) | =<br>(NS) |
| 83 | Steiner et al., 2019 [94] | Endurance athletes (n = 22) | Several venous blood parameters were assessed in ten male adolescent endurance athletes (five XC-skiers and five triathletes) at seven time points in 6-month intervals, resulting in a monitoring phase of 3 years. | =<br>(NS) | =<br>(NS) | - | =<br>(NS) | ↑<br>(3 y.) |

First, differences observed in [Hb] related to doping practises ranged from +10% following rhEPO doping [15] to −14% after blood withdrawal [16]. Otherwise, the range of increase or decrease in [Hb] was −5% due to certain pre-analytical conditions [17], +16% after acute exercise [18], −13% after prolonged exercise [19], −8% after exercise training [20], +9% after thermal acclimation [21], +4% after hypoxic training [22] and −12% after blood donation [23].

Subsequently, a large amplitude was observed in the relative changes in Ret% with increases up to +135% after rhEPO doping [24] and decreases of −38% observed after blood transfusions [16]. The range of increase or decrease in Ret% notwithstanding doping was +5% due to pre-analytical conditions [25], +25% after acute exercise [26], +63% after prolonged exercise [27], −44% after apnea training [28], +350% after hypoxic training [29] and +27% after blood donation [30].

Differences in the OFF-Score as a result of doping practises ranged from +18% after rhEPO doping [24] to −38% after blood transfusion [16]. The margin of increase or decrease in the OFF-Score related to other factors was −4% for pre-analytical conditions [31], −15% after prolonged exercise [32], −16% after exercise training [33], −5% after thermal acclimation [34] and +22% after hypoxic training [35].

Hct differences ranged from +10% after rhEPO intake [15] to −15% after blood transfusion [16]. The range of Hct variations related to other confounding factors was +4% [13] and −4% [17] for pre-analytical conditions, +12% after acute exercise [18], −8% after prolonged exercise [36], −7% after exercise training [20], −2% after thermal acclimation [37], +5% after hypoxic training [35] and +11% [38] and −11% [23] after blood donation.

Finally, PV increased by 14% after chronic xenon inhalation [39]. The range of increase or decrease in PV linked to other parameters was +4% for pre-analytical conditions [31], −20% after acute exercise [18], +24% after prolonged exercise [19], +16% after exercise training [20], +18% after thermal acclimation [40] and −14% after blood donation [38].

## 4 Discussion

Our study confirmed that the most obvious factor confounding the ABP biomarkers is blood doping. Doping protocols for erythropoiesis-stimulating substances (such as rhEPO injections) were generally structured with a treatment phase causing an increase in [Hb] and Ret% (ON-phase), followed by a reversal trend when treatment was stopped (OFF-phase) [24,41,42], although significant inter-individual variability was observed [41]. Following blood transfusions, an increase in [Hb] combined with a decrease in Ret% was generally reported after blood withdrawal while the opposite effect occurred during re-infusion [16,44]. In addition, chronic exposure to low doses of carbon monoxide was recently shown to positively influence erythropoiesis and alter markers sensitive to PV variations [46,95]. Conversely, repeated intake of desmopressin or chronic xenon inhalation induced haemodilution and decreased concentration-based biomarkers sensitive to PV shift [39,47,96]. However, micro-dosing doping schemes complicate the analysis of blood profiles due to limited haematological variations [45]. Indeed, the efficiency of the ABP appears significantly lower with low-volume autologous transfusion protocols due to several factors that may influence its sensitivity (e.g. timing of the sample collection) [45]. The current findings outline, for instance, the need for a strategy able to detect minor blood manipulations [97].

Numerous studies have highlighted specific blood variations as the result of pre-analytical factors including circadian modifications [25,48] and prevention strategies such as sodium intake [17], overhydration [31] or posture adjustments [13]. In the same way, a temporary [Hb] increase caused by PV reduction was usually observed following acute exercise [7,26,52,53] without affecting Ret% in most cases [51]. Similarly, acute exposure to extreme environmental conditions, such as heat [18,54–56], cold [57,58] or hypoxia [59], was reported to cause transitory PV shifts. Nevertheless, the strict WADA guidelines incorporate various pre-analytical precautions (e.g. reporting any exposure to hypoxia or extreme environment and other pathological conditions) to account for possible pre-analytical variations. However, other factors are not considered in the current model and may alter a proper interpretation of ABP profiles.

Prolonged exertion may affect an athlete’s blood values with temporary but delayed effects persisting for several days [60], and these effects may occur and persist independent of environmental conditions. A progressive decrease of [Hb] was observed during multiday events (e.g. cycling stage races) [19,32,61,66,67], although other studies observed a stabilisation over time [36,62] or even an increase towards the end of the competition [63]. The Ret% was mostly unaffected by participation in multiday events [32], although some variations (unrelated to an erythropoietic stimulation) were reported [27,60].

In contrast to transient variations caused by acute exercise, PV was shown to expand over a few weeks of endurance training, thereby reducing [Hb] [20,33,69]. Interestingly, multiple studies also reported a noticeable variation in Ret% after a few weeks of training [20,33,69]. Furthermore, acute and chronic training loads, the competition calendar and training periodisation were shown to significantly alter blood variables included in the ABP [70,75,76,98], with [Hb] decreases most likely during periods with the highest training loads [73]. In a recent 12-month longitudinal study of elite cyclists addressing the influence of training on ABP variables, no ATPF was observed, underlining the relative robustness of the ABP adaptive model in incorporating varying training loads [98].

Various environmental conditions (e.g. hypoxic or hot environments) are currently gaining popularity as additional training stimuli [99] with a putative effect on fluid balance [100]. PV increase represents one of the main physiological adaptations that occur during heat acclimation strategies [8,101], and this increase is, in turn, often linked to [Hb] reduction [37,40,77–79]. Although PV varies after heat acclimatisation, such variation may also occur without influencing ABP values [80], depending on the type of acclimatisation strategy and subjects involved. Another form of exposure to extreme environments is dry sauna bathing [102], which resulted in some heat acclimation with a significant PV expansion (up to ∼15%) [37,40]. A recent systematic review minimises the confounding risk of heat acclimation because no significant change in PV variations was reported after various acclimation protocols [103,104]. Conversely, several sessions of whole-body cryotherapy were proposed to decrease [Hb] [34,81].

Hypoxia is another environmental condition now widely used by athletes [105] where [Hb] may be either not [35,82] or only slightly affected [22] depending on the timing of blood sampling after *living high-training high* (LHTH) periods. Moreover, an increase in Ret% was reported during hypoxic exposure [82,83], while Ret% decreased upon return to a lower altitude [35]. Similar blood variations were reported following *living high-training low* (LHTL) protocols with, however, a more pronounced increase in [Hb] [84–87]. Variations due to hypoxic training observed immediately after altitude exposure were reported to persist three weeks after returning to sea level [35,84]. In addition, intermittent hypoxic exposure (IHE) or training (IHT) may positively influence Ret% [29]. Nevertheless, the rationale for the use of IHE for erythropoietic purposes in athletes is limited [106], and the response is inconsistent when hypoxic exposure is not prolonged [88,89]. Prolonged exposure to simulated or real altitude was reported to induce similar and prominent changes in total haemoglobin mass (Hb_mass_) [107,108]. Nevertheless, haematological variations following hypoxic training are contradictory [82,105] and not systematically reported [85,109], possibly due to initial fitness, initial fatigue or iron status [66,110]. It seems unlikely, then, that training in a hypoxic environment could lead to a misinterpretation of an athlete’s blood profiles [87] if duly reported in the doping control forms as requested by WADA.

Finally, specific individual characteristics were also shown to impact haematological components in athletes. Some haematological disorders were reported to alter [Hb], with these athletes exhibiting lower values than those of healthy athletes [90]. With some athletes suffering from haemochromatosis (a pathological condition requiring the withdrawal of large amounts of blood), [Hb] was decreased immediately after withdrawal [111] while a measurable increase in reticulocytes was sometimes delayed for a few days [30]. However, most haematological disorders observed in athletes were not associated with exceeding the limits of the model used to detect rhEPO [90]. Thus, the potential of these disorders to cause misinterpretation of the ABP profiles is limited. Finally, in contrast to Ret%, which is frequently higher among women [48,112], [Hb] is known to be higher in men than in women. Ret% was also shown to vary during the menstrual cycles of active women with lower values reported in the follicular phase [91]. Still, most of these variations remained within the individual ABP limits.

## 5 Conclusion

In conclusion, the effects of ABP confounders vary widely in amplitude and duration depending on the type of effector (i.e. doping, environmental condition, training or pre-analytical condition). Nevertheless, the absolute effects of the factors described in this review appear to be relatively limited when taken together. Furthermore, true and systematic effects of environmental conditions (i.e. heat acclimation or hypoxic training) on haematological biomarkers remain debatable due to significant differences in individual responses [104,105]. Studies investigating specific ABP confounders generally report variations within individual limits of the adaptive model [13,85,87,98]. The scientific level of ABP experts reviewing passports and their efforts to do so with the latter factors in mind can further limit misinterpretations of ABP profiles caused by the confounding factors identified in this review. Nevertheless, many authors have noted an important inter- and intra-variability concerning haematological biomarkers [112,113], highlighting the need to improve the sensitivity of the ABP while interpreting confounders carefully. The present review contributes to a detailed understanding of confounding factors that could affect ABP biomarkers. Our findings support the haematological module of the ABP as an efficient instrument to deter and indirectly detect doping, while further studies on the confounders that affect blood variables may contribute to improving the module and thus fighting doping more effectively.

## Data Availability

All data referred to in the manuscript is available on request to the corresponding author.

## Abbreviations

ABP: athlete biological passport
ADAMS: Anti-Doping Administration & Management System
APMU: Athlete Passport Management Unit
ATPF: atypical passport finding
[Hb]: haemoglobin concentration
Hb_mass_: haemoglobin mass
Hct: haematocrit
IHE: intermittent hypoxic exposure
IHT: intermittent hypoxic training
LHTH: living high-training high
LHTL: living high-training low
OFFs: OFF-Score
PV: plasma volume
Ret%: percentage of reticulocytes
WADA: World Anti-Doping Agency

## Declarations

### Ethics approval and Consent to participate

The present systematic review only includes data from existing and published studies so that no specific ethics approval was required

### Consent for publication

Not applicable

### Availability of data and materials

Not applicable

### Authors’ contribution

BK and RF conceived the project and designed the study. BK and RF contributed to the collection of data and interpreted the results. BK wrote the first draft of the manuscript. BK and RF contributed to revising the manuscript and expressed their approval of the final submitted version.

## Acknowledgements

Not applicable

## Authors’ information

Not applicable

## References

1. WADA. Athlete Biological Passport Operating Guidelines - version 7.1, June 2019. World Antidoping Agency. 2019. https://www.wada-ama.org/sites/default/files/resources/files/guidelines_abp_v71.pdf. 1 Oct 2020.

2. Saugy M, Leuenberger N. Antidoping: From health tests to the athlete biological passport. Drug Test Anal. 2020;12:621–28.

3. Sottas P-E, Robinson N, Saugy M. The Athlete’s Biological Passport and Indirect Markers of Blood Doping. Handb Exp Pharmacol. 2010;195:305–26.

4. Banfi G. Limits and pitfalls of Athlete’s Biological Passport. Clin Chem Lab Med. 2011;49:1417–21.

5. Sottas P-E, Baume N, Saudan C, Schweizer C, Kamber M, Saugy M. Bayesian detection of abnormal values in longitudinal biomarkers with an application to T/E ratio. Biostatistics. 2007;8:285–96.

6. Kargotich S, Goodman C, Keast D, Morton AR. The Influence of Exercise-Induced Plasma Volume Changes on the Interpretation of Biochemical Parameters Used for Monitoring Exercise, Training and Sport. Sports Med. 1998;26:101–17.

7. Lobigs LM, Sottas P-E, Bourdon PC, Nikolovski Z, El-Gingo M, Varamenti E, et al. A step towards removing plasma volume variance from the Athlete’s Biological Passport: The use of biomarkers to describe vascular volumes from a simple blood test. Drug Test Anal. 2018;10:294–300.

8. Sawka MN, Convertino VA, Eichner ER, Schnieder SM, Young AJ. Blood volume: importance and adaptations to exercise training, environmental stresses, and trauma/sickness. Med Sci Sports Exerc. 2000;32:332–48.

9. Sanchis-Gomar F, Martinez-Bello VE, Gomez-Cabrera MC, Viña J. Current limitations of the Athlete’s Biological Passport use in sports. Clin Chem Lab Med. 2011;49:1413–15.

10. WADA. Blood Analytical Requirements for the Athlete Biological Passport TD2010BAR 1.0. 1.06.2019;Version 1.0. World Antidoping Agency. 2009. https://www.wada-ama.org/sites/default/files/resources/files/guidelines_abp_v71.pdf. Accessed 1 Oct 2020.

11. WADA. Blood Sample Collection Guidelines. World Antidoping Agency. 2016. https://www.wada-ama.org/sites/default/files/resources/files/guidelines_blood_sample_collection_v4_0_2016_final_eng.pdf. Accessed 1 Oct 2020.

12. Kuuranne T, Saugy M, Baume N. Confounding factors and genetic polymorphism in the evaluation of individual steroid profiling. Br J Sports Med. 2014;48:848–55.

13. Astolfi T, Schumacher YO, Crettaz von Roten F, Saugy M, Faiss R. Does body position before and during blood sampling influence the Athlete Biological Passport variables? Int J Lab Hematol. 2020;42:61–7.

14. Wedin JO, Henriksson AE. The Influence of Floorball on Hematological Parameters: Consequences in Health Assessment and Antidoping Testing. J Sports Med. 2020;2020:1–6.

15. Haile DW, Durussel J, Mekonen W, Ongaro N, Anjila E, Mooses M, et al. Effects of EPO on Blood Parameters and Running Performance in Kenyan Athletes. Med Sci Sports Exerc. 2019;51:299–307.

16. Damsgaard R, Munch T, Mørkeberg J, Mortensen SP, Gonzàlez-Alonso J. Effects of blood withdrawal and reinfusion on biomarkers of erythropoiesis in humans: Implications for anti-doping strategies. Haematologica. 2006;91:1006–8.

17. Kuipers H, Brouwer T, Dubravcic-Simunjak S, Moran J, Mitchel D, Shobe J, et al. Hemoglobin and Hematocrit Values After Saline Infusion and Tourniquet. Int J Sports Med. 2005;26:405–8.

18. Diaz FJ, Bransford DR, Kobayashi K, Horvath SM, McMurray RG. Plasma volume changes during rest and exercise in different postures in a hot humid environment. J Appl Physiol. 1979;47:798–803.

19. Voss SC, Alsayrafi M, Bourdon P, Klodt F, Nonis D, Hopkins W, et al. Variability of Serum Markers of Erythropoiesis during 6 Days of Racing in Highly Trained Cyclists. Int J Sports Med. 2013;35:89–94.

20. Montero D, Breenfeldt-Andersen A, Oberholzer L, Haider T, Goetze JP, Meinild-Lundby A-K, et al. Erythropoiesis with endurance training: dynamics and mechanisms. Am J Physiol Regul Integr Comp Physiol. 2017;312:894–902.

21. Checinska-Maciejewska Z, Niepolski L, Checinska A, Korek E, Kolodziejczak B, Kopczynski Z, et al. Regular cold water swimming during winter time affects resting hematological parameters and serum erythropoietin. J Physiol Pharmacol. 2019;70:747–56.

22. Wachsmuth NB, Völzke C, Prommer N, Schmidt-Trucksäss A, Frese F, Spahl O, et al. The effects of classic altitude training on hemoglobin mass in swimmers. Eur J Appl Physiol. 2013;113:1199–211.

23. Meurrens J. Effect of Repeated Whole Blood Donations on Aerobic Capacity and Hemoglobin Mass in Moderately Trained Male Subjects: A Randomized Controlled Trial. Sports Med. 2016;2:12.

24. Bejder J, Aachmann-Andersen NJ, Bonne TC, Olsen NV, Nordsborg NB. Detection of erythropoietin misuse by the Athlete Biological Passport combined with reticulocyte percentage: Sensitivity of the adaptive model of the Athlete Biological Passport. Drug Test Anal. 2016;8:1049–55.

25. Sennels HP, Jørgensen HL, Hansen A-LS, Goetze JP, Fahrenkrug J. Diurnal variation of hematology parameters in healthy young males: The Bispebjerg study of diurnal variations. Scand J Clin Lab Invest. 2011;71:532–41.

26. Morici G, Zangla D, Santoro A, Pelosi E, Petrucci E, Gioia M, et al. Supramaximal exercise mobilizes hematopoietic progenitors and reticulocytes in athletes. Am J Physiol Regul Integr Comp Physiol. 2005;289:1496–503.

27. Fallon KE, Sivyer G, Sivyer K, Dare A. Changes in haematological parameters and iron metabolism associated with a 1600 kilometre ultramarathon. Br J Sports Med. 1999;33:27–31.

28. Revelli L, Vagnoni S, D’Amore A, Di Stasio E, Lombardi C, Storti G, et al. EPO Modulation in a 14-Days Undersea Scuba Dive. Int J Sports Med. 2013;34:856–60.

29. Kasperska A, Zembron-Lacny A. The effect of intermittent hypoxic exposure on erythropoietic response and hematological variables in elite athletes. Physiol Res. 2020;69:283–90.

30. Stangerup I, Kramp NL, Ziegler AK, Dela F, Magnussen K, Helge JW. Temporary impact of blood donation on physical performance and hematologic variables in women: blood donation and performance in women. Transfusion. 2017;57:1905–11.

31. Bejder J, Hoffmann MF, Ashenden MJ, Nordsborg NB, Karstoft K, Mørkeberg J. Acute hyperhydration reduces athlete biological passport OFF-hr score: Hyperhydration and athlete biological passport. Scand J Med Sci Sports. 2016;26:338–47.

32. Schumacher YO, Temme J, Bueltermann D, Schmid A, Berg A. The influence of exercise on serum markers of altered erythropoiesis and the indirect detection models of recombinant human erythropoietin abuse in athletes. Haematologica. 2003;88:712–4.

33. Bejder J, Andersen AB, Goetze JP, Aachmann-Andersen NJ, Nordsborg NB. Plasma volume reduction and hematological fluctuations in high-level athletes after an increased training load. Scand J Med Sci Sports. 2017;27:1605–15.

34. Lombardi G, Lanteri P, Porcelli S, Mauri C, Colombini A, Grasso D, et al. Hematological Profile and Martial Status in Rugby Players during Whole Body Cryostimulation. Young RC, editor. PLoS ONE. 2013;8:e55803.

35. Bonne TC, Lundby C, Lundby AK, Sander M, Bejder J, Nordsborg NB. Altitude training causes haematological fluctuations with relevance for the Athlete Biological Passport: Altitude training and the Athlete Biological Passport. Drug Test Anal. 2015;7:655–62.

36. Rama L, Minuzzi L, Carvalho H, Costa R, Teixeira A. Changes of Hematological Markers during a Multi-stage Ultra-marathon Competition in the Heat. Int J Sports Med. 2015;37:104–11.

37. Scoon GSM, Hopkins WG, Mayhew S, Cotter JD. Effect of post-exercise sauna bathing on the endurance performance of competitive male runners. J Sci Med Sport. 2007;10:259–62.

38. Ryan BJ, Goodrich JA, Schmidt WF, Stothard ER, Wright KP, Byrnes WC. Haemoglobin mass alterations in healthy humans following four-day head-down tilt bed rest. Exp Physiol. 2016;101:628–40.

39. Dias KA, Lawley JS, Gatterer H, Howden EJ, Sarma S, Cornwell WK, et al. Effect of acute and chronic xenon inhalation on erythropoietin, hematological parameters, and athletic performance. J Appl Physiol. 2019;127:1503–10.

40. Stanley J, Halliday A, D’Auria S, Buchheit M, Leicht AS. Effect of sauna-based heat acclimation on plasma volume and heart rate variability. Eur J Appl Physiol. 2015;115:785–94.

41. Bornø A, Aachmann-Andersen NJ, Munch-Andersen T, Hulston CJ, Lundby C. Screening for recombinant human erythropoietin using [Hb], reticulocytes, the OFFhr score, OFF z score and Hb z score: status of the Blood Passport. Eur J Appl Physiol. 2010;109:537–43.

42. Clark B, Woolford SM, Eastwood A, Sharpe K, Barnes PG, Gore CJ. Temporal changes in physiology and haematology in response to high- and micro-doses of recombinant human erythropoietin: Temporal responses to high- and micro-doses of rHuEPO. Drug Test Anal. 2017;9:1561–71.

43. Ashenden MJ, Gough CE, Garnham A, Gore CJ, Sharpe K. Current markers of the Athlete Blood Passport do not flag microdose EPO doping. Eur J Appl Physiol. 2011;111:2307–14.

44. Lamberti N, Finotti A, Gasparello J, Lampronti I, Zambon C, Cosenza LC, et al. Changes in hemoglobin profile reflect autologous blood transfusion misuse in sports. Intern Emerg Med. 2018;13:517–26.

45. Bejder J, Breenfeldt Andersen A, Solheim SA, Gybel-Brask M, Secher NH, Johansson PI, et al. Time Trial Performance Is Sensitive to Low-Volume Autologous Blood Transfusion. Med Sci Sports Exerc. 2019;51:692–700.

46. Schmidt WFJ, Hoffmeister T, Haupt S, Schwenke D, Wachsmuth NB, Byrnes WC. Chronic Exposure to Low Dose Carbon Monoxide Alters Hemoglobin Mass and VO2max. Med Sci Sports Exerc. 2020;52:1879–87.

47. Sanchis-Gomar F, Martinez-Bello VE, Nascimento AL, Perez-Quilis C, Garcia-Gimenez JL, Viña J, et al. Desmopresssin and Hemodilution: Implications in Doping. Int J Sports Med. 2010;31:5–9.

48. Robinson N, Saugy J, Schütz F, Faiss R, Baume N, Giraud S, et al. Worldwide distribution of blood values in elite track and field athletes: Biomarkers of altered erythropoiesis. Drug Test Anal. 2019;11:567–77.

49. Schumacher YO, Klodt F, Nonis D, Pottgiesser T, Alsayrafi M, Bourdon PC, et al. The impact of long-haul air travel on variables of the athlete’s biological passport. Int J Lab Hematol. 2012;34:641–7.

50. Alsaadi K, Voss SC, Kraiem S, Alwahaibi A, Alyazedi S, Dbes N, et al. The effect of fasting during Ramadan on parameters of the haematological and steroidal modules of the athletes biological passport – a pilot study. Drug Testing. 2015;7:1017–24.

51. Kuipers H, Dubravcic-Simunjak S, Moran J, Mitchell D, Shobe J, Sakai H, et al. Blood Testing in Sport: Hematological Profiling. Int J Sports Med. 2010;31:542–7.

52. Robinson N, Saugy M, Mangin P. Effects of exercise on the secondary blood markers commonly used to suspect erythropoietin doping. Clin Lab. 2003;49:57–62.

53. Miller GD, Teramoto M, Smeal SJ, Cushman D, Eichner D. Assessing serum albumin concentration following exercise-induced fluid shifts in the context of the athlete biological passport. Drug Test Anal. 2019;11:782–91.

54. Kenefick RW, Sollanek KJ, Charkoudian N, Sawka MN. Impact of skin temperature and hydration on plasma volume responses during exercise. J Appl Physiol. 2014;117:413–20.

55. Myhre LG, Robinson S. Fluid shifts during thermal stress with and without fluid replacement. J Appl Physiol. 1977;42:252–6.

56. Jimenez C, Melin B, Koulmann N, Allevard AM, Launay JC, Savourey G. Plasma volume changes during and after acute variations of body hydration level in humans. Eur J Appl Physiol. 1999;80:1–8.

57. Vogelaere P, Brasseur M, Quirion A, Leclercq R, Laurencelle L, Bekaert S. Hematological variations at rest and during maximal and submaximal exercise in a cold environment. Int J Biometeorol. 1990;34:1–14.

58. Vogelaere P, Savourey G, Deklunder G, Lecroart J, Brasseur M, Bekaert S, et al. Reversal of cold induced haemoconcentration. Europ J Appl Physiol. 1992;64:244–9.

59. Siebenmann C, Cathomen A, Hug M, Keiser S, Lundby AK, Hilty MP, et al. Hemoglobin mass and intravascular volume kinetics during and after exposure to 3,454-m altitude. J Appl Physiol. 2015;119:1194–201.

60. Miller GD, Beharry A, Teramoto M, Lai A, Willick SE, Eichner D. Hematological changes following an Ironman triathlon: An antidoping perspective. Drug Test Anal. 2019;11:1747–54.

61. Mørkeberg J, Belhage B, Damsgaard R. Changes in Blood Values in Elite Cyclist. Int J Sports Med. 2009;30:130–8.

62. Corsetti R, Lombardi G, Lanteri P, Colombini A, Graziani R, Banfi G. Haematological and iron metabolism parameters in professional cyclists during the Giro d’Italia 3-weeks stage race. Clin Chem Lab Med. 2012;50:949–56.

63. Lombardi G, Lanteri P, Fiorella PL, Simonetto L, Impellizzeri FM, Bonifazi M, et al. Comparison of the Hematological Profile of Elite Road Cyclists during the 2010 and 2012 GiroBio Ten-Day Stage Races and Relationships with Final Ranking. PLoS ONE. 2013;8:e63092.

64. Zappe DH, Tankersley CG, Meister TG, Kenny WL. Fluid Restriction Prior to Cycle Exercise: Effects on Plasma Volume and Plasma Proteins. Med Sci Sports Exerc. 1993;25:1225–30.

65. Gaebelein CJ, Senay LC. Influence of exercise type, hydration, and heat on plasma volume shifts in men. J Appl Physiol. 1980;49:119–23.

66. Garvican-Lewis LA, Schumacher YO, Clark SA, Christian R, Menaspà P, Plowman J, et al. Stage racing at altitude induces hemodilution despite an increase in hemoglobin mass. J Appl Physiol. 2014;117:463–72.

67. Schumacher YO, Garvican LA, Christian R, Lobigs LM, Qi J, Fan R, et al. High altitude, prolonged exercise, and the athlete biological passport: Athlete biological passport, altitude, and exercise. Drug Test Anal. 2015;7:48–55.

68. Spodaryk K. Haematological and iron-related parameters of male endurance and strength trained athletes. Europ J Appl Physiol. 1993;67:66–70.

69. Green HJ, Sutton JR, Coates G, Ali M, Jones S. Response of red cell and plasma volume to prolonged training in humans. J Appl Physiol. 1991;70:1810–5.

70. Banfi G, Del Fabbro M. Behaviour of reticulocyte counts and immature reticulocyte fraction during a competitive season in elite athletes of four different sports: reticulocytes in athletes. Int J Lab Hematol. 2007;29:127–31.

71. Abellan R, Ventura R, Pichini S, Palmi I, Bellver M, Olive R, et al. Effect of Physical Fitness and Endurance Exercise on Indirect Biomarkers of Recombinant Erythropoietin Misuse. Int J Sports Med. 2007;28:9–15.

72. Malcovati L, Pascutto C, Cazzola M. Hematologic passport for athletes competing in endurance sports: a feasibility study. Haematologica. 2003;88:570–81.

73. Banfi G, Del Fabbro M, Mauri C, Corsi MM, Melegati G. Haematological parameters in elite rugby players during a competitive season. Clin Lab Haematol. 2006;28:183–8.

74. Banfi G, Tavana R, Freschi M, Lundby C. Reticulocyte profile in top-level alpine skiers during four consecutive competitive seasons. Eur J Appl Physiol. 2010;109:561–8.

75. Mujika I, Goya A, Padilla S, Grijalba A, Gorostiaga E, Ibanez J. Physiological responses to a 6-d taper in middle-distance runners: influence of training intensity and volume. Med Sci Sports Exerc. 2000;32:511–7.

76. Mujika I, Goya A, Ruiz E, Grijalba A, Santisteban J, Padilla S. Physiological and Performance Responses to a 6-Day Taper in Middle-Distance Runners: Influence of Training Frequency. Int J Sports Med. 2002;23:367–73.

77. Costa RJS, Crockford MJ, Moore JP, Walsh NP. Heat acclimation responses of an ultra-endurance running group preparing for hot desert-based competition. Eur J Sport Sci. 2014;14:131–41.

78. Loeppky J. Plasma volume after heat acclimation: Variations due to season, fitness and methods of measurement. Acta Physiol Hung. 2015;102:282–92.

79. Tebeck ST, Buckley JD, Bellenger CR, Stanley J. Differing Physiological Adaptations Induced by Dry and Humid Short-Term Heat Acclimation. Int J Sports Physiol Perform. 2020;15:133–40.

80. Oberholzer L, Siebenmann C, Mikkelsen CJ, Junge N, Piil JF, Morris NB, et al. Hematological Adaptations to Prolonged Heat Acclimation in Endurance-Trained Males. Front Physiol. 2019;10:1379.

81. Banfi G, Krajewska M, Melegati G, Patacchini M. Effects of whole-body cryotherapy on haematological values in athletes. Br J Sports Med. 2008;42:858.

82. Garvican L, Martin D, Quod M, Stephens B, Sassi A, Gore C. Time course of the hemoglobin mass response to natural altitude training in elite endurance cyclists: Altitude training and hemoglobin mass. Scand J Med Sci Sports. 2012;22:95–103.

83. Ashenden MJ, Hahn AG, Martin DT, Logan P, Parisotto R, Gore CJ. A comparison of the physiological response to simulated altitude exposure and r-HuEpo administration. J Sports Sci. 2001;19:831–7.

84. Ashenden MJ, Gore CJ, Parisotto R, Sharpe K, Hopkins WG, Hahn AG. Effect of altitude on second-generation blood tests to detect erythropoietin abuse by athletes. Haematologica. 2003;88:1053–62.

85. Garvican-Lewis LA, Vuong VL, Govus AD, Schumacher YO, Hughes D, Lovell G, et al. Influence of combined iron supplementation and simulated hypoxia on the haematological module of the athlete biological passport. Drug Test Anal. 2018;10:731–41.

86. Neya M, Enoki T, Ohiwa N, Kawahara T, Gore CJ. Increased Hemoglobin Mass and VO2max With 10 h Nightly Simulated Altitude at 3000 m. Int J Sports Physiol Perform. 2013;8:366–72.

87. Voss SC, Al-Hamad K, Samsam W, Cherif A, Georgakopoulos C, Al Maadheed M, et al. A novel mixed living high training low intervention and the hematological module of the athlete biological passport. Drug Test Anal. 2020;12:323–30.

88. Abellan R, Ventura R, Remacha AF, Rodríguez FA, Pascual JA, Segura J. Intermittent hypoxia exposure in a hypobaric chamber and erythropoietin abuse interpretation. J Sports Sci. 2007;25:1241–50.

89. Sanchez AMJ, Borrani F. Effects of intermittent hypoxic training performed at high hypoxia level on exercise performance in highly trained runners. J Sports Sci. 2018;36:2045–52.

90. Parisotto R, Ashenden MJ, Gore CJ, Sharpe K, Hopkins W, Hahn AG. The effect of common hematologic abnormalities on the ability of blood models to detect erythropoietin abuse by athletes. Haematologica. 2003;88:931–40.

91. Mullen J, Bækken L, Bergström H, Björkhem Bergman L, Ericsson M, Ekström L. Fluctuations in hematological athlete biological passport biomarkers in relation to the menstrual cycle. Drug Test Anal. 2020;12:1229–40.

92. Lobigs LM. The Development Of Athlete-Specific Hematological Reference Intervals And The Effect Of Ethnicity On Key Hematological Markers. Qatar Foundation Annual Research Forum Proceedings. 2013;2013:BIOSP 07.

93. Steiner T, Wehrlin JP. Does Hemoglobin Mass Increase from Age 16 to 21 and 28 in Elite Endurance Athletes? Med Sci Sports Exerc. 2011;43:1735–43.

94. Steiner T, Maier T, Wehrlin JP. Effect of Endurance Training on Hemoglobin Mass and V·O2max in Male Adolescent Athletes. Med Sci Sports Exerc. 2019;51:912–9.

95. Sutehall S, Martin-Rincon M, Wang G, Shurlock J, Durussel J, Mooses M, et al. The Performance Effects of Microdose Recombinant Human Erythropoietin Administration and Carbon Monoxide Rebreathing. Curr Sports Med Rep. 2018;17:457–66.

96. Frampas C, Augsburger M, Varlet V. Xenon: From medical applications to doping uses. Toxicologie Analytique et Clinique. 2017;29:309–19.

97. Solheim SA, Bejder J, Breenfeldt Andersen A, Mørkeberg J, Nordsborg NB. Autologous Blood Transfusion Enhances Exercise Performance—Strength of the Evidence and Physiological Mechanisms. Sports Med - Open. 2019;5:30.

98. Astolfi T, Crettaz von Roten F, Kayser B, Saugy M, Faiss R. The influence of training load on hematological Athlete Biological Passport variables in elite cyclists [Prepress]. MedrXiv; 2020 Oct. Available from: http://medrxiv.org/lookup/doi/10.1101/2020.10.22.20213413

99. Baranauskas MN, Constantini K, Paris HL, Wiggins CC, Schlader ZJ, Chapman RF. Heat Versus Altitude Training for Endurance Performance at Sea Level. Exerc Sport Sci Rev. 2021;49:50–8.

100. Saunders PU, Garvican-Lewis LA, Chapman RF, Périard JD. Special Environments: Altitude and Heat. Int J Sport Nutr Exerc Metab. 2019;29:210–9.

101. Périard JD, Travers GJS, Racinais S, Sawka MN. Cardiovascular adaptations supporting human exercise-heat acclimation. Auton Neurosci. 2016;196:52–62.

102. Hussain J, Cohen M. Clinical Effects of Regular Dry Sauna Bathing: A Systematic Review. Evid Based Complement Alternat Med. 2018;2018:1–30.

103. Corbett J, Rendell RA, Massey HC, Costello JT, Tipton MJ. Inter-individual variation in the adaptive response to heat acclimation. J Therm Biol. 2018;74:29–36.

104. Rahimi GRM, Albanaqi AL, Van der Touw T, Smart NA. Physiological Responses to Heat Acclimation: A Systematic Review and Meta-Analysis of Randomized Controlled Trials. J Sports Sci Med. 2019;18:316–26.

105. Płoszczyca K, Langfort J, Czuba M. The Effects of Altitude Training on Erythropoietic Response and Hematological Variables in Adult Athletes: A Narrative Review. Front Physiol. 2018;9:375.

106. Faiss R, Girard O, Millet GP. Advancing hypoxic training in team sports: from intermittent hypoxic training to repeated sprint training in hypoxia: Table 1. Br J Sports Med. 2013;47:45–50.

107. Hauser A, Schmitt L, Troesch S, Saugy JJ, Cejuela-Anta R, Faiss R, et al. Similar Hemoglobin Mass Response in Hypobaric and Normobaric Hypoxia in Athletes. Med Sci Sports Exerc. 2016;48:734–41.

108. Saugy JJ, Schmitt L, Hauser A, Constantin G, Cejuela R, Faiss R, et al. Same Performance Changes after Live High-Train Low in Normobaric vs. Hypobaric Hypoxia. Front Physiol. 2016;7:138.

109. Siebenmann C, Robach P, Jacobs RA, Rasmussen P, Nordsborg N, Diaz V, et al. “Live high–train low” using normobaric hypoxia: a double-blinded, placebo-controlled study. J Appl Physiol. 2012;112:106–17.

110. Lobigs LM, Sharpe K, Garvican-Lewis LA, Gore CJ, Peeling P, Dawson B, et al. The athlete’s hematological response to hypoxia: A meta-analysis on the influence of altitude exposure on key biomarkers of erythropoiesis. Am J Hematol. 2018;93:74–83.

111. Johnson DM, Roberts J, Gordon D. The acute effects of whole blood donation on cardiorespiratory and haematological factors in exercise: A systematic review. PLoS ONE. 2019;14:e0215346.

112. Lombardi G, Colombini A, Lanteri P, Banfi G. Reticulocytes in sports medicine: an update. Adv Clin Chem. 2013;59:125–53.

113. Ashenden MJ, Lacoste A, Orhant E, Audran M, Sharpe K. Longitudinal Variation of Hemoglobin and Reticulocytes in Elite Rowers. Haematologica. 2004;89:1403–4.

